# The Effects of Cardiovascular Exercise on Corticospinal Excitability in People with Subacute Stroke

**DOI:** 10.64898/2025.12.02.25341471

**Authors:** Bernat De Las Heras, Lynden Rodrigues, Jacopo Cristini, Roya Khalili, Ada Tang, Janice J Eng, Joyce Fung, Marc Roig

## Abstract

Cardiovascular exercise (CE) shows promise for stroke recovery, partly by inducing neuroplasticity through excitatory neural signaling. While mechanisms are well-documented in animal models, the neurophysiological effects of CE in humans post-stroke—especially in the early subacute phase when the brain may be more responsive—remain unclear. In this study, 76 individuals within 3 months of their first-ever ischemic stroke were randomized to eight weeks of progressive CE using recumbent steppers plus standard care, or standard care alone. Corticospinal excitability (CSE) was assessed bilaterally using single and paired-pulse transcranial magnetic stimulation (TMS) at baseline, four weeks, and eight weeks. TMS was delivered at rest (chronic effect) and following a single high-intensity interval training (HIIT) session (acute effect). A single HIIT session at baseline significantly increased acute CSE in the contralesional hemisphere. However, despite significant improvements in cardiorespiratory fitness—indicating the effectiveness of the CE intervention—CE training did not lead to significant chronic or acute changes in CSE compared to standard care. This is the first study to investigate the effects of CE on CSE in early subacute stroke. Our findings indicate that while CE improves fitness levels during this critical period of recovery, it may have limited effects on CSE. **Trial Registration:** https://clinicaltrials.gov/study/NCT05076747.

## INTRODUCTION

Despite initial phases of neural damage and inflammation, the post-stroke brain exhibits a heightened yet time-limited state of neuroplasticity that contributes to recovery (Murphy and Corbett 2009). During this critical period, which spans about a month post-stroke in animal models, sensorimotor recovery is usually accompanied by profound structural and functional changes in the surviving neural systems, particularly in peri-infarct regions such as the primary motor cortex (M1). These changes include enhanced dendritic spine turnover, axonal sprouting, and cortical remapping, all tightly coupled to shifts in cellular excitability within cortical circuits (Carmichael 2003; Joy and Carmichael 2021; Krakauer et al. 2012).

Critically, this transient state of neural malleability can be harnessed through appropriate treatment interventions, with animal studies showing that early motor rehabilitation can lead to larger motor gains and neuroplastic changes compared to initiating rehabilitation at later stages (Biernaskie 2004; Yang et al. 2003). In humans, evidence has suggested a similar critical period within the first week to three months post-stroke, known as the early subacute phase (Bernhardt et al., 2017). Motor training interventions initiated during this period may lead to greater functional gains (Dromerick et al. 2021; Marzolini 2021). Hence, implementing motor rehabilitative treatments during early subacute stages of recovery may hold recovery potential.

Cardiovascular exercise (CE) is a core component of stroke rehabilitation due to its capacity to simultaneously improve functional, cardiorespiratory, and metabolic recovery outcomes (Billinger et al. 2014). Animal studies have demonstrated that functional recovery following CE can also be attributed to structural and functional changes in the nervous system (Ploughman and Kelly 2016), including upregulation of growth factors, neurogenesis, as well as enhanced synaptic plasticity (Austin et al. 2014; Ploughman et al. 2015), an event tightly regulated by cortical excitability (Clarkson and Carmichael 2009). However, unlike in animal models, the effects of CE on neuroplasticity mechanisms, particularly on brain excitability, remain largely unknown in people after stroke (Austin et al. 2014; Ploughman et al. 2015).

Given its capacity to assess the functional integrity of the corticospinal tract (CST), corticospinal excitability (CSE), assessed with transcranial magnetic stimulation (TMS), is one of the most extensively used biomarkers in stroke research (Dimyan and Cohen 2010; Talelli et al. 2006). When applied to the M1, TMS can activate a mixed population of inhibitory and facilitatory circuits, which depolarize to local and remote pyramidal tract neurons, enabling the quantification of distinct CSE measures (Rossini et al. 2015). Some of these measures have proven valuable for predicting motor recovery (Stinear et al. 2017) and assessing neurophysiological responses following motor rehabilitation treatments (Beaulieu and Milot 2018).

An increasing number of studies have investigated the impact of CE on CSE in post-stroke individuals. While findings are not always consistent, both acute and chronic changes have been reported in specific aspects of CSE following a single CE session and long-term training programs, respectively (De Las Heras et al. 2024). Recent studies have suggested modulatory effects of this intervention on CSE; however, they have focused on chronic recovery stages (> six months post-stroke) (Rodrigues et al. 2025), with no studies conducted during early subacute stages—thus neglecting earlier periods when corticospinal circuits may exhibit heightened responsiveness to neuromodulatory interventions. This randomized controlled trial (RCT) investigated, for the first time, the chronic and acute effects of CE on CSE in individuals during early subacute stages post-stroke.

## MATERIALS AND METHODS

In this two-arm single-site RCT (NCT05076747), participants were randomly assigned to either an 8-week CE training in addition to standard care or standard care alone **(Figure 1)**. Assessments occurred at baseline (T0), four weeks (T1), and eight weeks (T2). Each assessment comprised three experimental sessions 48 hours apart, covering clinical motor outcomes, cardiorespiratory fitness, and CSE measures. Participants were instructed not to engage in moderate- or high-intensity physical activity 24 hours before assessments. Enrollment occurred between June 2018 and July 2023. This trial followed the CONSORT (Consolidated Standards of Reporting Trials) guidelines. The local ethics review board approved the study (Centre de Recherche de Readaptation du Montréal, CRIR-1265-0817), and all participants provided written informed consent.

**Figure 1.**
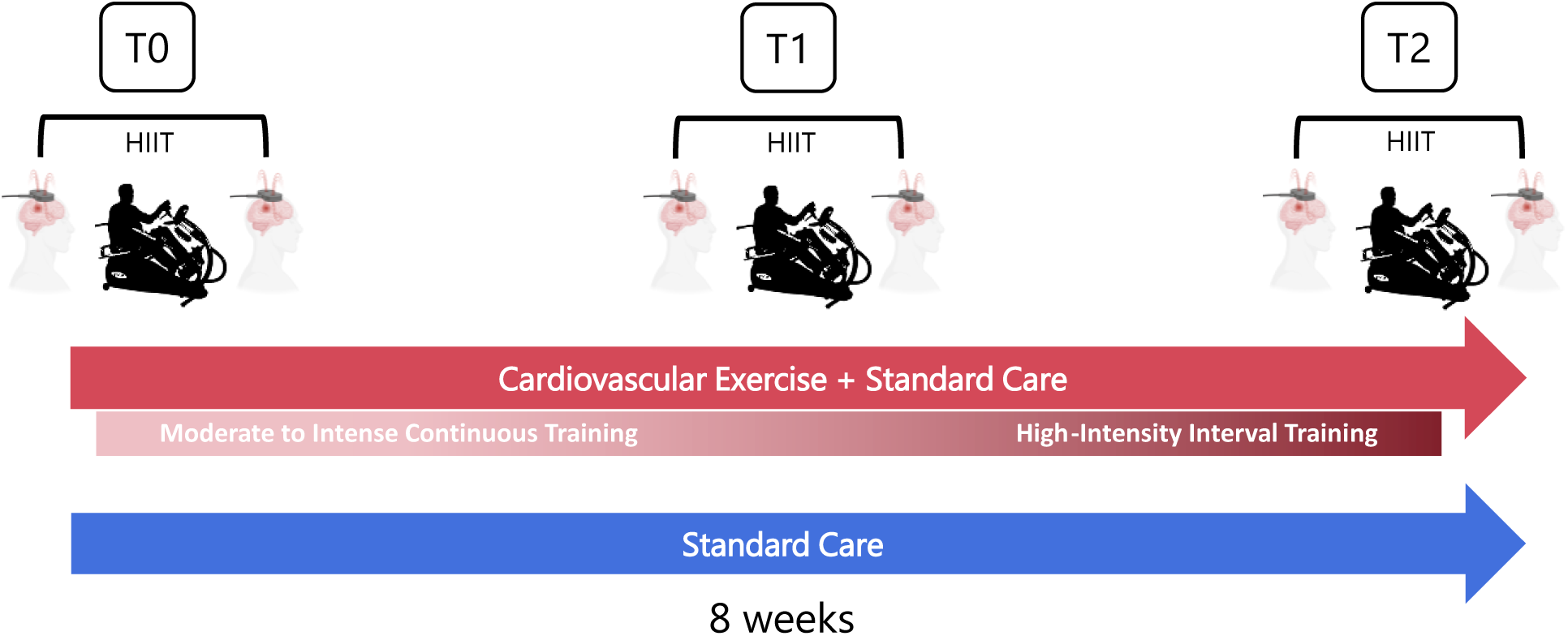
Study design: TMS evaluations were performed at baseline (T0), four weeks (T1), and eight weeks (T2). To measure the chronic effects of CE training on CSE measures, TMS was applied at rest at each time point. Acute CSE changes were determined by measuring the difference between pre- and post- a 15-minute HIIT session (post-HIIT–pre-HIIT), with loads individually adjusted based on a previous GXT. CE: cardiovascular exercise, CSE: corticospinal excitability, GXT: graded exercise test, HIIT: high-intensity interval training, TMS: transcranial magne c stimulation.

### Participants

We included participants aged 40-80 years with first-ever ischemic stroke within the early subacute stages (7 days-3 months post-stroke) (Bernhardt et al. 2017). Eligibility required no other musculoskeletal or neurological conditions, sufficient cognitive/communicative capacity to perform the exercise protocols and follow instructions safely, and no TMS contraindications (Rossini et al. 2015). Exclusion criteria were hemorrhagic stroke, cognitive impairment and/or aphasia affecting informed consent, absolute contraindications to exercise (Ferguson 2014), or concurrent enrollment in another exercise program. Stroke lesions were classified as cortical, subcortical, or cerebellar based on clinical reports from radiologists who analyzed CT/MRI scans within two days of the event.

## ASSESSMENTS

### Cardiorespiratory Fitness

Peak oxygen uptake (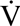O_2_peak in mL.Kg^−1^.min^−1^) was measured during a symptom-limited graded exercise test (GXT) (Ferguson 2014). A validated protocol (Billinger et al. 2008) was performed on a whole-body recumbent stepper (NuStep T4r, Michigan, USA) with resistance increasing every 2 minutes. Heart rate (HR) was measured continuously, while blood pressure and rate of perceived exertion (RPE 0-10; (Borg 1970)) were taken every 2 minutes. The GXT also provided maximal HR (HR_max_) expressed as beats per minute (bpm) and peak power output (PPO) expressed as Watts (W). 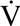O_2_peak was the highest recorded oxygen uptake. Tests were terminated upon volitional exhaustion, absolute termination criteria (Fletcher et al. 2013), or inability to maintain ≥80 steps per minute after two warnings.

### Clinical Motor Outcomes

Upper-limb motor impairment changes were assessed with the Upper-Limb Fugl-Meyer Assessment (UL-FMA), with higher scores indicating lower impairment (Lin JH 2004). Upper-limb function were assessed with the Box and Block Test (BBT) (Lin et al. 2010), in which participants used their affected hand to move as many blocks as possible across a partition within one minute.

### Transcranial Magnetic Stimulation

TMS assessments at T0, T1, T2 were conducted at a similar time of day to minimize circadian variations. Head, hand, and arm positions, including seat recline and arm placement angles, were recorded at T0 and replicated at subsequent sessions to ensure consistency. Maximal voluntary contraction (MVC) of each hand was assessed using a hand-grip force transducer controlled by a custom LabView script (National Instruments, Austin, TX, USA). Participants completed two 3-second squeezes per hand with 30-second rests, receiving visual force feedback; the highest MVC per hand was recorded.

Using neuronavigation (Brainsight, Rogue Research Inc., Montreal, QC, Canada), we first co-registered the patient’s head to a standard MRI template to guide TMS targeting. A stereotaxic, grid-guided mapping procedure was used to elicit motor-evoked potentials (MEPs) from M1, with the “hot spot” defined as the coil position producing the largest and most consistent MEPs. TMS was applied through a 70-mm coil with a Magstim BiStim stimulator (Magstim, Whitland, Wales, UK), oriented 45⁰ posterior to the midsagittal line (Rossini et al. 2015), targeting the first dorsal interosseous (FDI) muscle representation in both the ipsilesional and contralesional hemispheres. Electromyographic activity was recorded via two surface electrodes placed ∼1 cm apart over the FDI, acquired at 2000 Hz through a CED Micro1401-4 unit controlled by Signal software (CED, Cambridge, UK), with a gain of 300 and band-pass filtered (10–500 Hz) (Ostadan et al. 2016).

### Corticospinal Excitability

To investigate chronic and acute CSE responses to CE, single- and paired-pulse TMS protocols were performed before and 10 minutes after a 15-minute standardized high-intensity interval exercise (HIIT) session, except for resting motor threshold (MT), which was measured only at rest (Hallett 2007). To minimize potential repetitive TMS effects on MEP amplitude, pulses were delivered every 5 seconds, with 30 stimulations per hemisphere for each CSE measure. The HIIT protocol began and ended with 3 minutes of warm-up and cool-down at 15% PPO, and consisted of six 30-second blocks of high-intensity at 90% PPO (total 3 minutes), interspersed with six 1-minute bouts at 30% PPO (total 6 minutes) (Boyne et al. 2015). Participants were instructed to maintain a minimum cadence of 80 steps/min (Nepveu et al. 2017), and intensity was continuously monitored through Watts, HR, and RPE.

*<u>Resting motor threshold:</u>* the resting MT was defined as the minimum stimulation intensity required to elicit MEPs of >0.05 mV in at least 10 of 20 trials (Rossini et al. 2015). Expressed as a percentage of stimulator output, resting MT reflects the neural membrane excitability of cortical axons, with higher resting MT representing lower CSE (Talelli et al. 2006).

*<u>MEP amplitude:</u>* MEP amplitudes were assessed at rest and during a sustained muscle contraction at 20% MVC. To assess active excitability, the LabView MVC script provided visual feedback to help participants maintain the target force. MEP amplitude was quantified as the average peak-to-peak value of responses elicited at 120% resting MT (Tunovic et al. 2014). This measure reflects the excitability of cortical and spinal projections regulated by both excitatory (glutamate) and inhibitory (γ-aminobutyric acid, GABA) circuits (Ziemann et al. 2015), with larger MEP amplitudes reflecting higher CSE. *<u>Cortical silent period (CSP)</u>*: The CSP reflects inhibitory activity modulated by GABA_B_ receptors, with longer CSP reflecting greater inhibition (Rossini et al. 2015). CSP was calculated from the EMG baseline amplitude recorded 200 ms pre-stimulation, measured from the end of the MEP to the return of voluntary activity—defined as an increase >2 SD above the mean baseline amplitude (Nepveu et al. 2017).

*<u>Intracortical facilitation and short intracortical inhibition:</u>* Intracortical facilitation (ICF) provides information on facilitation mediated by N-methyl-d-aspartate (NMDA) and glutamatergic neural activity, while short intracortical inhibition (SICI) reflects inhibition mediated by phasic GABA_A_-related signaling (Ziemann et al. 2015). Both were measured with a paired-pulse TMS protocol in which a conditioning pulse (80% resting MT) was followed by a suprathreshold pulse (120% resting MT) delivered after 10 (ICF) and 2 (SICI) milliseconds (ms), respectively. The amplitude of the conditioned MEP was normalized to the unconditioned MEP amplitudes at 120% RMT, expressed as a percentage, with values >100% indicating higher facilitation or reduced inhibition.

### Interventions

#### Cardiovascular Exercise

The CE + standard care group completed 24 CE training sessions over 8 weeks (3×/week, with ∼48 hours between sessions). Training comprised 4 weeks of progressive moderate-to-vigorous intensity continuous training (MICT) followed by 4 weeks of progressive HIIT on a whole-body recumbent stepper **(Figure 1)**. Each session included 2.5 minutes of warm-up and cool-down at 35% PPO, with the main training component performed at the targeted intensity. Blood pressure was measured pre- and post-session. Heart rate and power output were continuously monitored, while RPE (0-10; (Borg 1970)) was recorded every 5 minutes. Training variables—including average %HRmax, %PPO, total steps, and average RPE—were calculated to quantify internal and external workloads **(Table 2)**.

#### Moderate-to-vigorous Continuous Training (weeks 1-4)

MICT has been typically employed as standard CE training in stroke rehabilitation (Billinger et al. 2014). Intensities were determined using the PPO associated with VO_2_peak during the GXT at T0 and increased weekly by 5% from 65% to 80% PPO to promote adaptations. Session durations progressed from 20 to 35 minutes.

#### High-intensity Interval Training (weeks 5-8)

HIIT was prescribed using the PPO from the T1 GXT and consisted of 8 x 60-second high-intensity intervals (starting at 85% PPO and progressing 5% weekly to 100%) interspersed with 7 x 60-second low-intensity intervals at 35% PPO, totaling 20 minutes per session. This 60:60 interval ratio supports sustained high intensity in stroke patients (Boyne et al. 2015). To minimize abrupt BP changes, workload was gradually increased 15 seconds before each high-intensity interval.

### Standard Care Program

Standard care, delivered at the same center as the intervention and prescribed by the stroke clinical unit, included routine health monitoring by physicians and nursing staff, physiotherapy, occupational therapy, and speech therapy. Session content, duration, and frequency were individualized, with each session lasting 45 minutes. To examine potential differences between groups in standard care, the type and number of standard care sessions received by each patient were recorded throughout the study **(Table 1)**.

**Table 1.** Baseline demographic and clinical outcomes. AC, anticoagulant; ACE, angiotensin-converting Enzyme; AP, antiplatelet; BB, beta-blocker; BBT, box and block Test; BMI, body mass index; CCI, Charlson comorbidity index; F, female; UL-FMA, upper-limb Fugl-Meyer assessment; M, male; MET, metabolic equivalent of task; MoCA, Montreal cognitive assessment; MVC, maximal voluntary contraction; NIHSS, national institutes of health stroke scale; PSY, psychoactive; SNP, single nucleotide polymorphism; STA, statin. Values are presented as mean ± standard deviation (SD) unless otherwise specified.

|  | **CE + Standard**<br>**Care**<br>**(n= 48)** | **Standard Care**<br>**(n= 28)** |
| --- | --- | --- |
| **Age (years)** | 63±11.39 | 65.35±8.68 |
| **Sex (F/M)** | 11/37 | 10/18 |
| **BMI** | 27±3.46 | 26.52±4.01 |
| **Time since stroke (days)** | 68.12±22.07 | 58.75±24.03 |
| **Lesion location (%)** |  |  |
| **Cortical** | 36 | 36 |
| **Subcortical** | 54 | 57 |
| **Cerebellar** | 10 | 7 |
| **NIHSS (0-42)** | 2.02±2.20 | 1.92±1.92 |
| **MoCA (0-30)** | 24.14±4.85 | 23.21±3.8 |
| **UL-FMA (0-66)** | 56.20±10.24 | 59.14±8.22 |
| **BBT<sub>affected</sub> (blocks/min)** | 46.8±13.25 | 48.10±12.65 |
| **MVC<sub>affected</sub>** | 0.72±0.35 | 0.66±0.27 |
| **Cardiorespiratory fitness (VO<sub>2peak</sub>, mL.Kg<sup>-1</sup>.min<sup>-1</sup>)** | 17.88±5.49 | 18.12±5.58 |
| **CCI (Age-adjusted)** | 4.57±1.83 | 4.57±1.66 |
| **Walking aid dependence (%)** | 15 | 13 |
| **Smoking history (%)** |  |  |
| **Non-smoker** | 52 | 43 |
| **Former smoker** | 40 | 56 |
| **Current smoker** | 8 | 1 |
| **Medications (n)** | 5.07±2.58 | 5±2.22 |
| **Classification (%)** |  |  |
| **AC** | 60 | 53 |
| **ACE** | 31 | 42 |
| **AP** | 46 | 60 |
| **BB** | 35 | 25 |
| **PSY** | 33 | 28 |
| **STA** | 79 | 100 |
| **Therapy sessions (n)** |  |  |
| **Physiotherapy** | 8.87±8.21 | 6.59±5.78 |
| **Occupational Therapy** | 11.5±8.26 | 7.45±5.98 |
| **Speech Therapy** | 5.02±8.81 | 2.15±5.48 |
| **Physical activity (METS hr/day)** | 8.37±4.99 | 9.26±6.44 |

### Statistical Analysis

Analyses followed a prespecified intention-to-treat approach. Normality of variables was inspected via plots and histograms and confirmed with the Shapiro-Wilk test. Baseline differences between groups were assessed using t-tests or Wilcoxon tests. Influential observations were identified via quantile range outliers (tail quantile = 0.1, Q = 3). Linear mixed models (LMM) evaluated group differences over time (T0-T1-T2) for clinical motor outcomes, cardiorespiratory fitness, and CSE measures. Group differences for each CSE measure were analyzed separately for both ipsilesional and contralesional hemispheres. Each model included either acute or chronic CSE measures as the dependent variable, with timepoint, group, and their interaction as fixed effects, and age, sex, handgrip MVC, and stroke severity as covariates. Participants were treated as a random effect to account for individual differences at baseline. An AR(1) covariance structure was used based on Bayesian Information Criterion, log-likelihood ratio tests, and the data’s temporal dependence. Assumptions for linear models, including normality and homoscedasticity of residuals, were examined for all the variables in the model. Associations between CSE measures and changes in fitness and motor outcomes were investigated using standard least-squares multivariate linear regression, adjusted for the same covariates and are reported in supplementary files. Multicollinearity between predictor variables was assessed with the variance inflation factor with a threshold of ≤5, indicating unacceptable multicollinearity (De Havenon et al. 2021). Analyses were performed in JMP 17 (SAS Institute, Cary, NC) with significance set at p < 0.05.

## RESULTS

**Table 1** displays the participants’ characteristics and clinical information at baseline. The trial flow, including dropouts, is detailed in **Figure 2**. No significant differences were observed at T0 between groups in terms of age, sex, body mass index (BMI), time since stroke, lesion location, stroke severity, cognitive status, upper-limb impairment and function, pre-existing comorbidities, walking aid dependence, smoking history, and the average number of prescribed medications **(Table 1).** The amount of standard care, including physiotherapy, occupational therapy, and speech therapy, as well as levels of physical activity outside the rehabilitation center (measured with the PASIPD), were similar between groups from T0 to T2. All participants assigned to the CE+standard care group who completed the study attended all 24 CE sessions.

**Figure 2.**
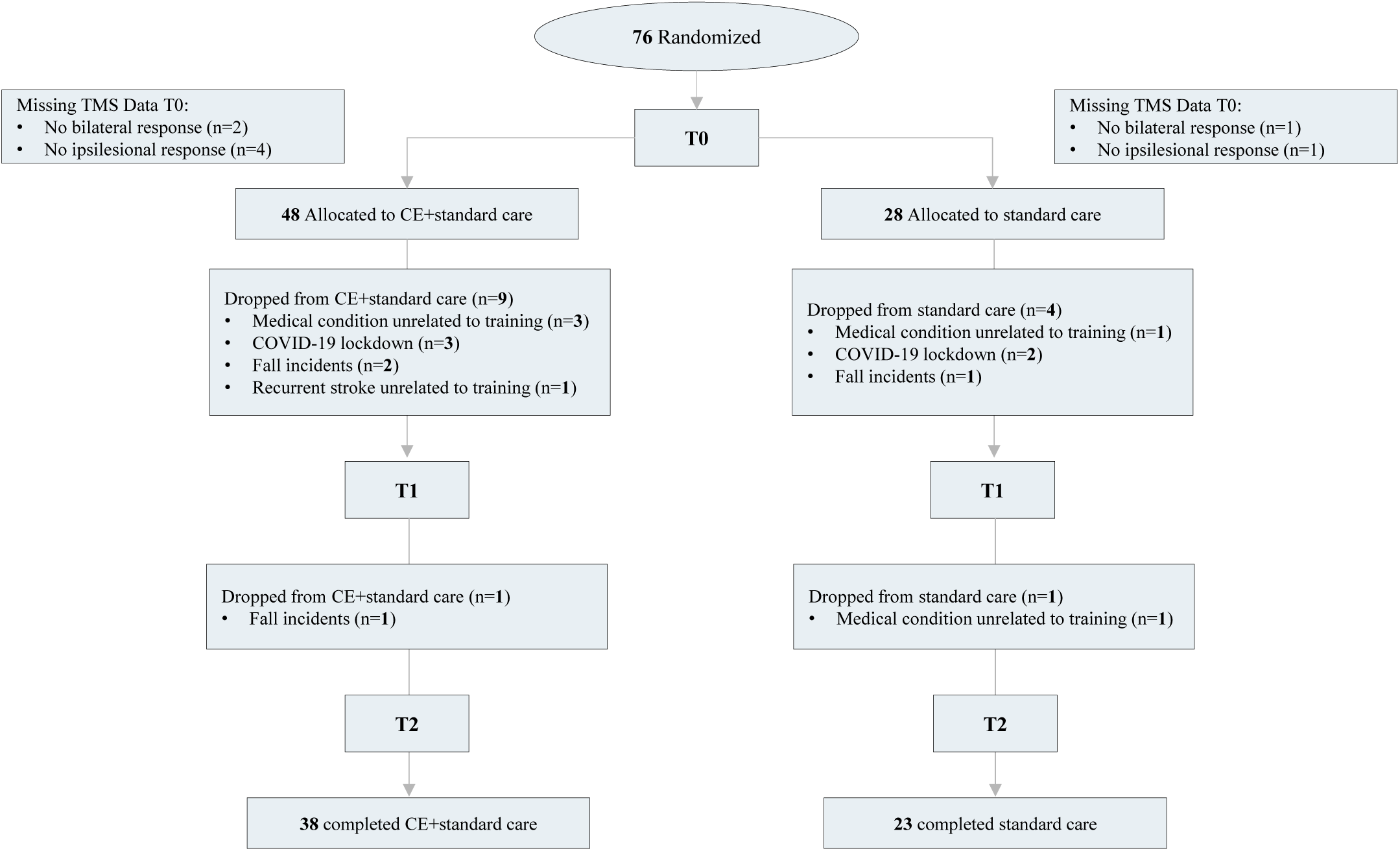
Flow chart of the Randomized Controlled Trial. CE: cardiovascular exercise, n: number of participants, TMS: transcranial magnetic stimulation, T0: baseline, T1: four weeks, T2: eight weeks

### Cardiorespiratory Fitness And Clinical Motor Outcomes

No significant differences in cardiorespiratory fitness were observed between groups at baseline (T0). At T0, all participants obtained an average VO_2_peak of 18.43±5.63 mL.Kg^−1^.min^−1^. There was a significant effect of Time (F(2,78)=16.76, p≤.0001), and a significant Time x Group interaction (F(2,78)=13.46, p≤.0001) in cardiorespiratory fitness. While the standard care group showed no significant change in VO_2_peak from T0 to T2 (+0.27 mL.Kg^−1^.min^−1^, 95% confidence interval [CI] −2.19 to 1.64, p=.998), the CE+standard care group showed significant 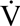O_2_peak improvements, with an increase of +2.76 mL.Kg^−1^.min^−1^ (95% CI: 1.58 to 3.93, p≤.0001) at T1 during MICT, further increasing by +1.64 mL.Kg^−1^.min^−1^ (95% CI: 0.45 to 2.82, P≤.0001) at T2 following HIIT, for a total gain of +4.43 mL.Kg^−1^.min^−1^ from T0 (95% CI: 2.97 to 5.82, P≤.0001). There was a significant effect of Time on upper-limb motor impairment and function, as measured by the UL-FMA (F(2, 99)=15.61, p≤.0001) and BBT (F(2, 116)=15.73, p≤.0001), indicating improvements across both groups. No significant Time x Group interaction was observed for either measure (UL-FMA: F(2, 99)=1.04, p=.355; BBT: F(2, 116)=0.22, p=.801). Changes in each measure for both groups are detailed in **Supplementary Table 1**.

**Table 2.**
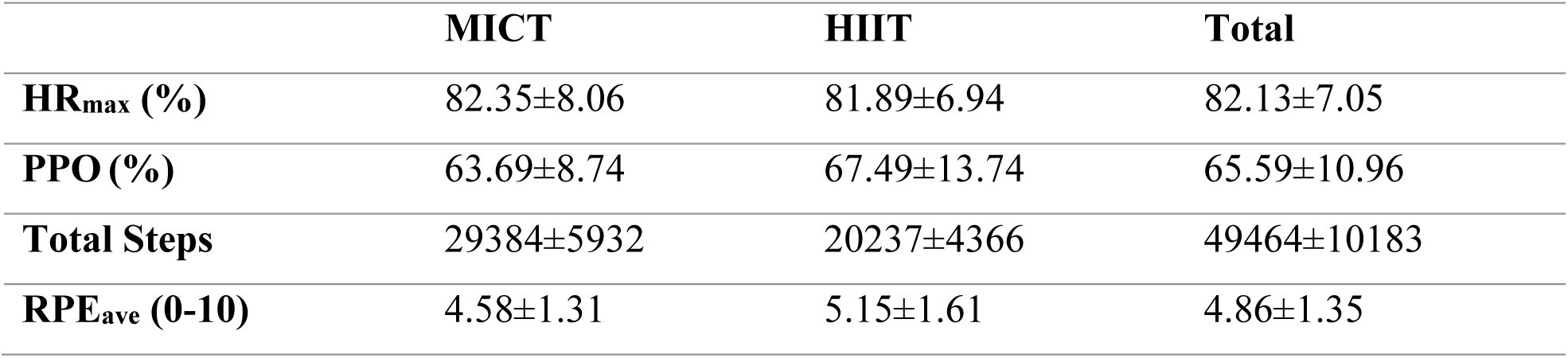
Internal and external training workloads for the CE group. CE group’s average internal and external training loads during both MICT and HIIT periods, including the warm-up and cool-down phases of each session. The average percentages of HRmax and PPO achieved during both MICT and HIIT periods were calculated based on VO2peak values during the GXT at T0 and T1, respectively. Values are presented as mean and SD. CE, cardiovascular exercise; HIIT, high-intensity interval training; HR, heart rate; MICT, moderate-to-vigorous Continuous Training; PPO, peak power output; RPE, rate of perceived exertion.

### Cardiovascular Exercise Effects On Corticospinal Excitability

The total percentage of TMS trials removed during the analysis was 0.04%. CSE measures were obtained from all participants except 3, whose MEPs could not be elicited on either hemisphere **(Figure 2)**. For 5 participants with no ipsilesional response, only contralesional hemisphere data were used. Data from all participants were included to measure CSE responses at T0, and an intention-to-treat approach was used for those who were assessed at least at T1. No statistically significant differences were observed between groups for any CSE measure at T0. When comparing ipsilesional and contralesional hemispheres, CSP showed significant differences, with greater prolongation ipsilesionally (F(1, 291.3)=53.63, p≤.0001), suggesting increased inhibition. These differences remained significant across timepoints and were consistent across both groups after adjusting for handgrip MVC.

No significant effects of Time or Time x Group interaction were found for chronic CSE changes (T0-T2) in either hemisphere (**Figure 3, Supplementary Table 2**). Similarly, no significant acute effects of Time or Time x Group interaction were identified for any CSE measures over time in either ipsilesional or contralesional hemispheres **(Figure 4, Supplementary Table 3)**. Chronic and acute changes for each CSE measure for both groups are detailed in **Supplementary Tables 2 and 3**, respectively.

**Figure 3.**
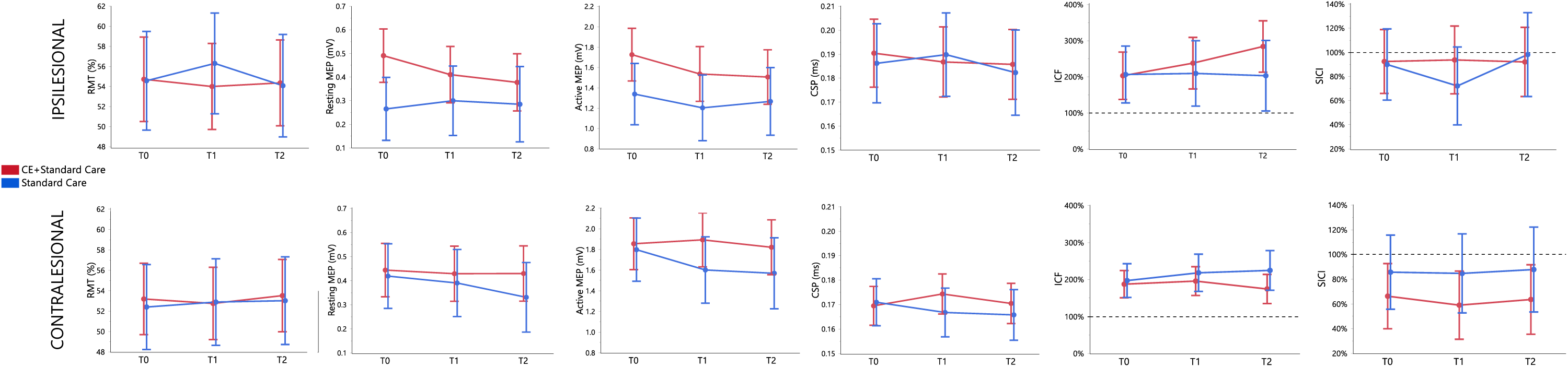
Chronic CSE changes in CE+standard care and standard care groups through time points (T0,T1,T2). Note that in ICF and SICI conditioned MEP amplitude is normalized to the unconditioned resting MEP amplitude with values >100% indicating higher facilitation and lower inhibition, respectively. Data is presented as least squares means and error bars are standard error of the means. CSP, cortical silent period; ICF, intracortical facilitation; MEP, motor evoked potential; ms, millisecond; mV, millivolt; SICI, short-intracortical inhibition.

**Figure 4.**
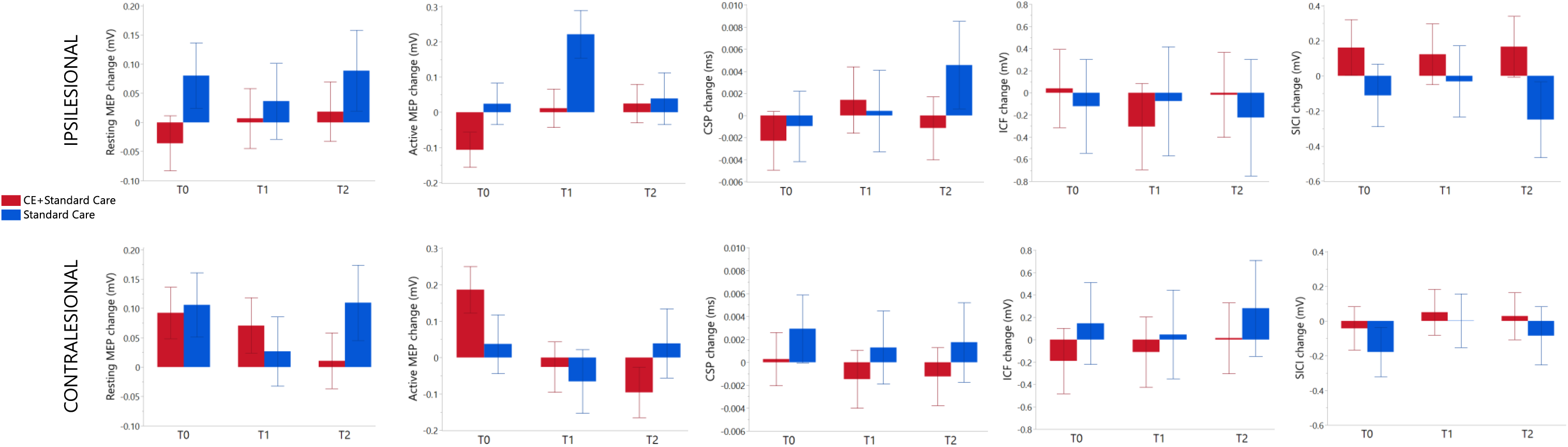
Acute CSE changes following a single HIIT session in CE+standard care and standard care groups through time points (T0,T1,T2). Data is presented as least squares means and error bars are standard error of the mean. CSP, cortical silent period; ICF, intracortical facilitation; MEP, motor evoked potential; ms, millisecond; mV, millivolt; SICI, short-intracortical inhibition.

An analysis combining both groups at T0 (n=74) to investigate the acute response to exercise before participants were randomized to the different arms of the study revealed significant increases in resting (F(1,70) = 7.77, p=0.006) and active (F(1,70) = 7.43, p=0.008) MEP amplitudes on the contralesional hemisphere following a 15-minute HIIT session, with no significant changes on the ipsilesional hemisphere **(Figure 5, Supplementary Table 4)**.

**Figure 5.**
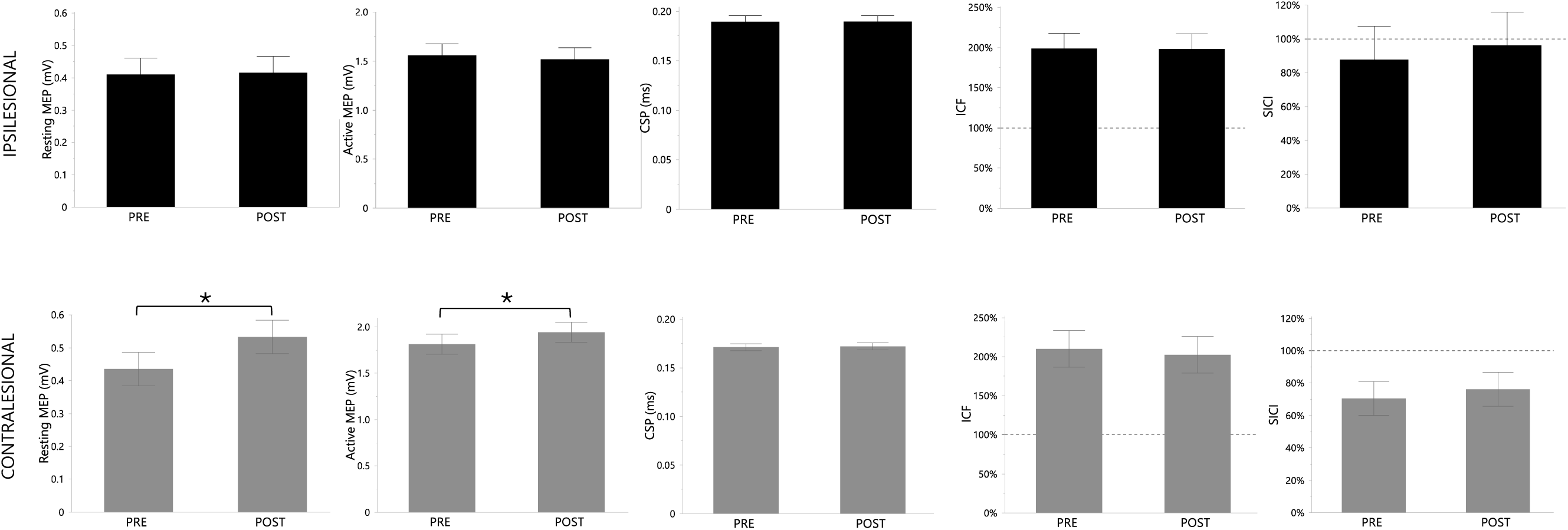
Acute CSE changes (PRE-POST) in response to a HIIT session at T0 (n=73). Note that in ICF and SICI conditioned MEP amplitude is normalized to the unconditioned resting MEP amplitude with values >100% indicating higher facilitation and lower inhibition, respectively. Data is presented as least squares means and error bars are standard error of the means. CSP, cortical silent period; ICF, intracortical facilitation; MEP, motor evoked potential; ms, millisecond; mV, millivolt; SICI, short-intracortical inhibition.

## DISCUSSION

In addition to enhancing cardiorespiratory and metabolic outcomes, CE has been proposed as a potential intervention to support stroke recovery by promoting neural adaptations that facilitate neural repair, such as changes in CSE (De Las Heras et al. 2024). This aligns with evidence from animal models showing that exercise-related motor recovery is linked to neuroplastic events modulated by cortical excitability, particularly during early post-stroke stages, when the brain may be more responsive to treatment (Biernaskie 2004). In our study, no significant effects were observed on chronic CSE responses following eight weeks of CE training **(Figure 3)**. These findings contrast with pre-clinical evidence indicating increases in synaptic excitability-associated markers, such as synaptogenesis and dendritic branching after repetitive CE (Ploughman et al. 2015), as well as with human data from three studies in the late-subacute (3-6 months) and chronic stages (>6 months) that reported long-term CSE increases following 4-12 weeks of CE training (Rodrigues et al. 2025; Yang et al. 2010; Yen et al. 2008). However, given the small sample sizes of most these studies, the lack of workload monitoring, the absence of exercise tests to assess cardiorespiratory fitness improvements, or the lack of a non-exercise control group, it remains unclear whether these CSE changes were directly attributable to the training stimulus and adaptations from the CE intervention (De Las Heras et al. 2024).

Our findings suggest that the lack of significant effects on CSE following CE training was not due to the intervention’s inability to improve cardiorespiratory fitness. In fact, compared to the control group receiving standard care alone, our progressive 8-week CE training program significantly increased cardiorespiratory fitness, with average VO_2_peak improvements of 4.43±3.24 mL.Kg^−1^.min^−1^. These improvements exceed previously reported values in subacute stroke populations undergoing high-intensity CE training interventions (+1.46 mL.Kg^−1^.min^−1^) (Luo et al. 2020), as well as the minimal clinically important difference (MCID) for 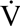O_2_peak (+3.5 mL.Kg^−1^.min^−1^). These substantial improvements observed in cardiorespiratory fitness, together with the lack of significant associations between 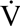O_2_peak increases and CSE in our multivariate linear regression analysis (**Supplementary Table 6**), suggest that our findings are unlikely to be attributed to an insufficient training stimulus. These results are consistent with the only CE study to date examining chronic CSE responses in neurotypical individuals, which revealed no significant CSE changes following six weeks of intensive cycling training, despite notable improvements in cardiorespiratory fitness (Nicolini et al. 2019). Together, this evidence suggests that while CE training confers significant cardiovascular benefits, its impact on CSE appears to be limited, at least during the early subacute stages of recovery.

While long-term interventions are essential to determine lasting changes in neural networks and their impact on recovery, a single exercise session has been widely used as a scientific paradigm to gain mechanistic insights into the neurophysiological responses elicited by CE (Nepveu et al. 2017). Studies using TMS in neurotypical individuals have demonstrated that a single session of CE can transiently modulate specific CSE parameters, with higher intensities evoking larger increases (Singh and Staines 2015). Notably, in some studies (Kuo et al. 2023; Ostadan et al. 2016; Stavrinos and Coxon 2017), but not all (Mang et al. 2014; Morris et al. 2020), these acute CSE modulations have been associated with motor skill learning, suggesting a potential mechanism for motor gains (Robertson and Takacs 2017). Furthermore, several studies in both rodents and neurotypical humans have indicated that long-term CE interventions can amplify the acute neurobiological response to a single CE session in certain neuroplasticity biomarkers such as brain-derived neurotrophic factor (Berchtold et al. 2005; Ikegami et al. 2024; Szuhany et al. 2015), as well as CSE (Lulic et al. 2017), supporting the notion of adaptive neurophysiological changes following CE training.

In our study, acute responses to CE were measured immediately following a single 15-minute HIIT protocol at baseline (T0) prior to group allocation, as well as at 4 weeks (T1) and 8 weeks (T2) in both groups to evaluate the potential effects of CE training on acute CSE responses. Regarding the long-term effects of CE training on the acute response, no significant changes in any CSE markers were observed following a 15-minute HIIT session at the 4-week (T1) or 8-week (T2) time points compared to standard care (**Figure 4)**. However, when combining data from both groups at baseline (T0), we observed significant acute increases in CSE within the contralesional hemisphere, as indicated by resting and active MEP amplitudes, while no changes were noted in the ipsilesional hemisphere **(Figure 5, Supplementary Table 4)**.

These findings demonstrate, for the first time, that certain aspects of CSE can be modulated by a single session of high-intensity exercise during the early-subacute stage of post-stroke recovery. This contrasts with studies in chronic stroke populations, which have reported acute CSE increases in the ipsilesional—but not contralesional—hemisphere following a single high-intensity CE session, both when performed alone (Sivaramakrishnan and Subramanian 2023) and following a 6-month long-term CE intervention (Forrester et al. 2006). Specifically, in individuals at the chronic stage of recovery, MEP latencies, MTs, and MEP amplitudes have been shown to respond to short CE bouts, with higher intensities evoking more pronounced CSE increases ipsilesionally (Abraha et al. 2018; Boyne et al. 2019; Li et al. 2019).

Conversely, the absence of acute changes in intracortical excitability measures, such as SICI and ICF, observed in our study aligns with prior research on patients in the chronic recovery stages (Abraha et al. 2018; Boyne et al. 2019; Li et al. 2019; Nepveu et al. 2017) but contrasts with findings in neurotypical populations, which consistently report increases in ICF and decreases in SICI after a single CE bout (Neva et al. 2017; Singh et al. 2014; Smith et al. 2014; Stavrinos and Coxon 2017; Yamazaki et al. 2019). Although direct comparisons between stroke and neurotypical populations are still lacking, these findings suggest a limited capacity for CE alone to modulate the facilitatory-inhibitory excitatory balance in stroke patients, a required precursor for neuroplastic changes supporting recovery (Joy and Carmichael 2021).

One of the possible explanations for the limited CSE changes following CE training observed in our study, and for the discrepancies between findings from subacute and chronic stages, may lie in the time-dependent neurobiological processes that characterize post-stroke recovery (Dobkin and Carmichael 2016). During the initial days to weeks post-stroke, neural connections are disrupted, resulting in diminished responses to afferent inputs in areas surrounding the lesion, causing an overall state of hypo-excitability (Carmichael 2016). This phenomenon is observed in animals but also in humans, where the ipsilesional hemisphere often exhibits reduced or even absent CSE, particularly during the acute and early subacute recovery phases (McDonnell and Stinear 2017). One of the key factors contributing to this reduced responsiveness is the increased inhibitory activity commonly observed during these early recovery stages. This period is characterized by an approximately 50% increase in GABAergic tone in areas adjacent to the lesion, primarily through tonic extrasynaptic GABA signaling (Cramer 2008). While this inhibition serves as a neuroprotective mechanism against excitotoxic signaling during acute stages (Green AR 2000; Lyden and Hedges 1992), its prolonged presence can reduce the propensity of neurons to fire in response to excitatory stimuli, hindering essential neuroplastic processes for recovery (Alia et al. 2016; Carmichael 2012). Indeed, pre-clinical studies reducing tonic GABA signaling—either pharmacologically or genetically—during early stages can promote motor recovery by restoring excitability in neural circuits (Alia et al. 2016; Clarkson et al. 2010; Orfila et al. 2019; Wang et al. 2018). Therefore, the absence of significant acute and chronic CSE changes in response to CE observed in our study, particularly in the ipsilesional hemisphere, could be partly attributed to a persistent inhibitory activity characteristic of the subacute phase. This interpretation aligns with a recent study from our group, in which 12 weeks of CE elicited ipsilesional CSE modulation in individuals with chronic stroke [median (IQR) = 20.25 (15.4) months post-stroke] (Rodrigues et al. 2025), underscoring the complex and evolving nature of the brain’s excitability response across different stages of recovery.

Supporting this hypothesis, our findings indicated a significant increase in inhibition as evidenced by one of our TMS measures. Specifically, ipsilesional CSP—but not SICI—remained significantly elevated compared to the contralesional hemisphere at all time points (T0, T1, T2), suggesting increased GABA_B_ receptor-mediated activity during subacute recovery stages. These findings align with previous evidence indicating increased ipsilesional GABA_B_, but not GABA_A_, synaptic activity in subacute post-stroke individuals (Cirillo et al. 2020). This discrepancy in GABAergic systems may arise because tonic extrasynaptic inhibition, rather than synaptic (phasic) inhibition, appears to be increased early after stroke (Clarkson et al. 2010), a molecular event undetectable through the 2-ms interstimulus interval SICI protocol used in this study (Stagg et al. 2011) but that could be indirectly captured through CSP (Connelly et al. 2013). Based on these findings, CSP could have captured elevated extrasynaptic inhibition in the affected corticomotor pathway, potentially serving as a mechanism explaining the limited responsiveness to CE during early subacute stages. Future studies should aim to validate this hypothesis using methods capable of measuring extracellular neurotransmitter levels, such as magnetic resonance spectroscopy (Coxon et al. 2018; Stagg et al. 2011).

In addition to these unique neurophysiological events, another factor that could have influenced our findings is the specificity or lack thereof the intervention (Kleim JA 2008). Evidence from animal and human studies suggests that unskilled, repetitive movement activity alone, without a motor learning component, does not induce significant lasting changes in corticospinal motor circuits (Kleim et al. 2002; Perez et al. 2006; Plautz et al. 2000). A similar principle has been observed in stroke motor rehabilitation, where nonspecific movement alone does not appear to elicit the same degree of neural and motor recovery changes as when is paired with targeted, intensive, goal-oriented training (Biernaskie and Corbett 2001; Debow et al. 2003; Jeffers and Corbett 2018; Maldonado et al. 2008; Marin et al. 2003; Ploughman et al. 2007). Cardiovascular exercise promotes a neurochemical milieu in the brain in ways that is supportive of neuroplasticity and overall brain function (Voss et al. 2013). However, evidence from animal models on early post-stroke stages shows that unspecific exercise can promote neuroplasticity and support functional recovery approaching pre-stroke levels but only when paired with skilled motor rehabilitation (Jeffers and Corbett 2018; Ploughman et al. 2007). Unlike rehabilitative interventions such as gait or upper-limb motor training, which tend to be task-specific and based on motor learning principles, our intervention was primarily designed to induce physiological and neurophysiological adaptations through a whole-body exercise using recumbent steppers, rather than improving a specific task (Krakauer 2006; Winstein et al. 2014). While we demonstrated that this mode of CE can be effective to improve cardiovascular fitness in stroke populations across the recovery continuum (De Las Heras et al. 2025b; Moncion et al. 2024), it may lack the specificity to induce significant neural changes in corticospinal motor circuits, particularly during early subacute stages, when inhibitory systems are likely upregulated. Future studies investigating the potential synergetic effects of combining CE training with goal-oriented motor training on CSE in subacute phases of recovery are warranted.

## LIMITATIONS

One of the main limitations of this study was the absence of raw neuroimaging data, which would have allowed for more precise lesion localization, particularly regarding the extent of CST damage. Transcranial magnetic stimulation offers a highly focused spatial resolution (1-2 cm) for assessing CSE from the CST (Dimyan and Cohen 2010; Talelli et al. 2006). Recent evidence indicates that lesion location can strongly influence CSE patterns, especially when subacortical descending corticospinal pathways are affected (De Las Heras et al. 2025a). Quantitatively controlling for CST integrity could have provided a more nuanced understanding of individual responses to CE. Although sensitivity analyses across lesion subtypes (cortical, subcortical, cerebellar) showed no significant effect on CSE **(Supplementary Table 5-6)**, variability in CST damage may still have influenced the observed effects on CSE. Future studies should integrate precise neuroimaging measures of CST lesion load (e.g., anisotropy, diffusion weighted imaging) alongside TMS assessments to better understand how CSE is modulated by CE interventions in stroke patients, where inter-subject variability is common (Feng et al. 2015).

Another limitation was the relatively low disability levels in our sample, which may restrict the generalizability of our findings to individuals with more severe impairments. While obtaining complete CSE measures from those with significant CST disruption can be challenging or even impossible (Stinear et al. 2006), evidence indicates that individuals with severe lesions may exhibit distinct neuroplasticity and CSE patterns (Buetefisch et al. 2023; Thickbroom et al. 2015). Future studies could address this limitation by using multimodal neuroimaging to characterize neural activity and neurotransmitter dynamics in individuals without elicitable MEPs (Stagg et al. 2011).

## CONCLUSION

We investigated, for the first time, the acute and chronic effects of CE in stroke patients during the early subacute phase of recovery. While a single HIIT session at baseline produced significant increases in contralesional hemisphere CSE, CE training did not elicit significant acute or chronic long-term changes in CSE, despite substantial gains in cardiorespiratory fitness. Factors such as increased inhibitory cortical activity during early stages of stroke recovery, lack of intervention specificity, and individual patient variability may have influenced these results. Overall, our findings support CE as an effective strategy to enhance cardiovascular fitness in the subacute phase of stroke recovery, but do not provide evidence that it promotes sustained CSE modulation at this stage.

## Data Availability

All data produced in the present study are available upon reasonable request to the authors

## AUTHOR CONTRIBUTIONS

Bernat De Las Heras: Data curation; Formal analysis; Investigation; Methodology; Project administration; Writing—original draft; and Writing—review & editing. Lynden Rodrigues: Data curation; Investigation; and Writing—review & editing. Jacopo Cristini: Data curation; Investigation; and Writing—review & editing. Roya Khalili: Data curation; Investigation; and Writing—review & editing. Ada Tang: Conceptualization; Funding acquisition; Supervision; and Writing—review & editing. Janice J Eng: Conceptualization; Funding acquisition; Methodology; and Writing—review & editing. Joyce Fung: Methodology and Writing—review & editing. Marc Roig: Conceptualization; Data curation; Formal analysis; Funding acquisition; Investigation; Methodology; Project administration; Resources; Software; Supervision; and Writing— review & editing.

## REFERENCES

Abraha B et al. 2018. A bout of high intensity interval training lengthened nerve conduction latency to the non-exercised affected limb in chronic stroke. Front Physiol. 9(827. https://www.ncbi.nlm.nih.gov/pubmed/30013489. doi:10.3389/fphys.2018.00827.

Alia C et al. 2016. Reducing gabaa-mediated inhibition improves forelimb motor function after focal cortical stroke in mice. Sci Rep. 6(1):37823. https://dx.doi.org/10.1038/srep37823. doi:10.1038/srep37823.

Austin MW, Ploughman M, Glynn L, Corbett D. 2014. Aerobic exercise effects on neuroprotection and brain repair following stroke: A systematic review and perspective. Neurosci Res. 87(8–15. https://dx.doi.org/10.1016/j.neures.2014.06.007. doi:10.1016/j.neures.2014.06.007.

Beaulieu L-D, Milot M-H. 2018. Changes in transcranial magnetic stimulation outcome measures in response to upper-limb physical training in stroke: A systematic review of randomized controlled trials. Ann Phys Rehabil Med. 61(4):224–234. https://dx.doi.org/10.1016/j.rehab.2017.04.003. doi:10.1016/j.rehab.2017.04.003.

Berchtold NC, Chinn G, Chou M, Kesslak JP, Cotman CW. 2005. Exercise primes a molecular memory for brain-derived neurotrophic factor protein induction in the rat hippocampus. Neuroscience. 133(3):853–861. https://www.ncbi.nlm.nih.gov/pubmed/15896913. doi:10.1016/j.neuroscience.2005.03.026.

Bernhardt J et al. 2017. Agreed definitions and a shared vision for new standards in stroke recovery research: The stroke recovery and rehabilitation roundtable taskforce. Int J Stroke. 12(5):444–450. https://www.ncbi.nlm.nih.gov/pubmed/28697708. doi:10.1177/1747493017711816.

Biernaskie J. 2004. Efficacy of rehabilitative experience declines with time after focal ischemic brain injury. 24(5):1245–1254. https://dx.doi.org/10.1523/JNEUROSCI.3834-03.2004. doi:10.1523/jneurosci.3834-03.2004.

Biernaskie J, Corbett D. 2001. Enriched rehabilitative training promotes improved forelimb motor function and enhanced dendritic growth after focal ischemic injury. The Journal of Neuroscience. 21(14):5272–5280. https://dx.doi.org/10.1523/jneurosci.21-14-05272.2001. doi:10.1523/jneurosci.21-14-05272.2001.

Billinger SA et al. 2014. Physical activity and exercise recommendations for stroke survivors. Stroke. 45(8):2532–2553. https://dx.doi.org/10.1161/str.0000000000000022. doi:10.1161/str.0000000000000022.

Billinger SA, Tseng BY, Kluding PM. 2008. Modified total-body recumbent stepper exercise test for assessing peak oxygen consumption in people with chronic stroke. 88(10):1188–1195. https://dx.doi.org/10.2522/ptj.20080072. doi:10.2522/ptj.20080072.

Borg. 1970. Perceived exertion as an indicator of somatic stress. Scand J Rehabil Med. 2(92–98.

Boyne P et al. 2015. Within-session responses to high-intensity interval training in chronic stroke. Med Sci Sports Exerc. 47(3):476–484. https://www.ncbi.nlm.nih.gov/pubmed/24977698. doi:10.1249/MSS.0000000000000427.

Boyne P et al. 2019. Exercise intensity affects acute neurotrophic and neurophysiological responses poststroke. J Appl Physiol. 126(2):431–443. https://dx.doi.org/10.1152/japplphysiol.00594.2018. doi:10.1152/japplphysiol.00594.2018.

Buetefisch CM et al. 2023. Stroke lesion volume and injury to motor cortex output determines extent of contralesional motor cortex reorganization. Neurorehabil Neural Repair.15459683231152816. https://www.ncbi.nlm.nih.gov/pubmed/36786394. doi:10.1177/15459683231152816.

Carmichael ST. 2016. The 3 rs of stroke biology: Radial, relayed, and regenerative. Neurotherapeutics. 13(2):348–359. https://dx.doi.org/10.1007/s13311-015-0408-0. doi:10.1007/s13311-015-0408-0.

Carmichael ST. 2012. Brain excitability in stroke: The yin and yang of stroke progression. Arch Neurol. 69(2):161–167. https://www.ncbi.nlm.nih.gov/pubmed/21987395. doi:10.1001/archneurol.2011.1175.

Carmichael ST. 2003. Plasticity of cortical projections after stroke. Neuroscientist. 9(1):64–75. https://www.ncbi.nlm.nih.gov/pubmed/12580341. doi:10.1177/1073858402239592.

Cirillo J et al. 2020. Neurochemical balance and inhibition at the subacute stage after stroke. J Neurophysiol. 123(5):1775–1790. https://www.ncbi.nlm.nih.gov/pubmed/32186435. doi:10.1152/jn.00561.2019.

Clarkson AN, Carmichael ST. 2009. Cortical excitability and post-stroke recovery. Biochem Soc Trans. 37(Pt 6):1412–1414. https://www.ncbi.nlm.nih.gov/pubmed/19909287. doi:10.1042/BST0371412.

Clarkson AN, Huang BS, Macisaac SE, Mody I, Carmichael ST. 2010. Reducing excessive gaba-mediated tonic inhibition promotes functional recovery after stroke. Nature. 468(7321):305–309. https://dx.doi.org/10.1038/nature09511. doi:10.1038/nature09511.

Connelly WM et al. 2013. Gaba breceptors regulate extrasynaptic gaba areceptors. The Journal of Neuroscience. 33(9):3780–3785. https://dx.doi.org/10.1523/jneurosci.4989-12.2013. doi:10.1523/jneurosci.4989-12.2013.

Coxon JP et al. 2018. Gaba concentration in sensorimotor cortex following high-intensity exercise and relationship to lactate levels. The Journal of Physiology. 596(4):691–702. https://dx.doi.org/10.1113/jp274660. doi:10.1113/jp274660.

Cramer SC. 2008. Repairing the human brain after stroke: I. Mechanisms of spontaneous recovery. Ann Neurol. 63(3):272–287. https://dx.doi.org/10.1002/ana.21393. doi:10.1002/ana.21393.

De Havenon A et al. 2021. Effect of adjusting for baseline stroke severity in the national inpatient sample. Stroke. 52(11). https://dx.doi.org/10.1161/strokeaha.121.035112. doi:10.1161/strokeaha.121.035112.

De Las Heras B et al. 2025a. Lesion location changes the association between brain excitability and the performance of a short-term visuomotor adaptation task post-stroke. Brain Communications. https://dx.doi.org/10.1093/braincomms/fcaf430. doi:10.1093/braincomms/fcaf430.

De Las Heras B et al. 2024. Measuring neuroplasticity in response to cardiovascular exercise in people with stroke: A critical perspective. Neurorehabil Neural Repair.15459683231223513. https://www.ncbi.nlm.nih.gov/pubmed/38291890. doi:10.1177/15459683231223513.

De Las Heras B et al. 2025b. Investigating the acute and chronic effects of cardiovascular exercise on brain-derived neurotrophic factor in early subacute stroke. Neurorehabil Neural Repair.15459683251342150. https://www.ncbi.nlm.nih.gov/pubmed/40462267. doi:10.1177/15459683251342150.

Debow SB, Davies MLA, Clarke HL, Colbourne F. 2003. Constraint-induced movement therapy and rehabilitation exercises lessen motor deficits and volume of brain injury after striatal hemorrhagic stroke in rats. Stroke. 34(4):1021–1026. https://dx.doi.org/10.1161/01.str.0000063374.89732.9f. doi:10.1161/01.str.0000063374.89732.9f.

Dimyan MA, Cohen LG. 2010. Contribution of transcranial magnetic stimulation to the understanding of functional recovery mechanisms after stroke. Neurorehabil Neural Repair. 24(2):125–135. http://mcgill.on.worldcat.org/atoztitles/link?sid=OVID:medline&id=pmid:19767591&id=doi:10.1177%2F1545968309345270&issn=1545-9683&isbn=&volume=24&issue=2&spage=125&pages=125-35&date=2010&title=Neurorehabilitation+%26+Neural+Repair&atitle=Contribution+of+transcranial+magnetic+stimulation+to+the+understanding+of+functional+recovery+mechanisms+after+stroke.&aulast=Dimyan.

Dobkin BH, Carmichael ST. 2016. The specific requirements of neural repair trials for stroke. Neurorehabilitation and Neural Repair. 30(5):470–478. https://dx.doi.org/10.1177/1545968315604400. doi:10.1177/1545968315604400.

Dromerick AW et al. 2021. Critical period after stroke study (cpass): A phase ii clinical trial testing an optimal time for motor recovery after stroke in humans. Proc Natl Acad Sci U S A. 118(39). https://www.ncbi.nlm.nih.gov/pubmed/34544853. doi:10.1073/pnas.2026676118.

Feng W et al. 2015. Corticospinal tract lesion load: An imaging biomarker for stroke motor outcomes. Ann Neurol. 78(6):860–870. https://dx.doi.org/10.1002/ana.24510. doi:10.1002/ana.24510.

Ferguson B. 2014. Acsm’s guidelines for exercise testing and prescription 9th ed. J Can Chiropr Assoc. 58(3):328.

Fletcher GF et al. 2013. Exercise standards for testing and training. Circulation. 128(8):873–934. https://dx.doi.org/10.1161/cir.0b013e31829b5b44. doi:10.1161/cir.0b013e31829b5b44.

Forrester LW, Hanley DF, Macko RF. 2006. Effects of treadmill exercise on transcranial magnetic stimulation-induced excitability to quadriceps after stroke. Arch Phys Med Rehabil. 87(2):229–234. https://proxy.library.mcgill.ca/login?url=http%3A%2F%2Fovidsp.ovid.com/ovidweb.cgi?T=JS&CSC=Y&NEWS=N&PAGE=fulltext&D=med6&AN=16442977 http://mcgill.on.worldcat.org/atoztitles/link?sid=OVID:medline&id=pmid:16442977&id=doi:10.1016%2Fj.apmr.2005.10.016&issn=0003-9993&isbn=&volume=87&issue=2&spage=229&pages=229-34&date=2006&title=Archives+of+Physical+Medicine+%26+Rehabilitation&atitle=Effects+of+treadmill+exercise+on+transcranial+magnetic+stimulation-induced+excitability+to+quadriceps+after+stroke.&aulast=Forrester.

Green AR HA, Jackson DM. 2000 Gaba potentiation: A logical pharmacological approach for the treatment of acute ischaemic stroke. Neuropharmacology. 10(9):1483–1494. doi:10.1016/s0028-3908(99)00233-6.

Hallett M. 2007. Transcranial magnetic stimulation: A primer. Neuron. 55(2):187–199. https://dx.doi.org/10.1016/j.neuron.2007.06.026. doi:10.1016/j.neuron.2007.06.026.

Ikegami R et al. 2024. In vivo bioluminescence imaging revealed the change of the time window of bdnf expression in the brain elicited by a single bout of exercise following repeated exercise. Neurosci Lett. 834(137830. https://www.ncbi.nlm.nih.gov/pubmed/38788795. doi:10.1016/j.neulet.2024.137830.

Jeffers MS, Corbett D. 2018. Synergistic effects of enriched environment and task-specific reach training on poststroke recovery of motor function. Stroke. 49(6):1496–1503. https://dx.doi.org/10.1161/strokeaha.118.020814. doi:10.1161/strokeaha.118.020814.

Joy MT, Carmichael ST. 2021. Encouraging an excitable brain state: Mechanisms of brain repair in stroke. Nature Reviews Neuroscience. 22(1):38–53. https://dx.doi.org/10.1038/s41583-020-00396-7. doi:10.1038/s41583-020-00396-7.

Kleim JA, Cooper NR, Vandenberg PM. 2002. Exercise induces angiogenesis but does not alter movement representations within rat motor cortex. Brain Res. 934(1):1–6. https://dx.doi.org/10.1016/s0006-8993(02)02239-4. doi:10.1016/s0006-8993(02)02239-4.

Kleim JA JT. 2008 Principles of experience-dependent neural plasticity: Implications for rehabilitation after brain damage. J Speech Lang Hear Res. 51(S225-239. doi:10.1044/1092-4388(2008/018).

Krakauer JW. 2006 Motor learning: Its relevance to stroke recovery and neurorehabilitation. Curr Opin Neurol 19(84–90.

Krakauer JW, Carmichael ST, Corbett D, Wittenberg GF. 2012. Getting neurorehabilitation right: What can be learned from animal models? Neurorehabil Neural Repair. 26(8):923–931. https://www.ncbi.nlm.nih.gov/pubmed/22466792. doi:10.1177/1545968312440745.

Kuo HI, Hsieh MH, Lin YT, Kuo MF, Nitsche MA. 2023. A single bout of aerobic exercise modulates motor learning performance and cortical excitability in humans. Int J Clin Health Psychol. 23(1):100333. https://www.ncbi.nlm.nih.gov/pubmed/36168600. doi:10.1016/j.ijchp.2022.100333.

Li X, Charalambous CC, Reisman DS, Morton SM. 2019. A short bout of high-intensity exercise alters ipsilesional motor cortical excitability post-stroke. Top Stroke Rehabil. 26(6):405–411. http://mcgill.on.worldcat.org/atoztitles/link?sid=OVID:medline&id=pmid:31144609&id=doi:10.1080%2F10749357.2019.1623458&issn=1074-9357&isbn=&volume=26&issue=6&spage=405&pages=405-411&date=2019&title=Topics+in+Stroke+Rehabilitation&atitle=A+short+bout+of+high-intensity+exercise+alters+ipsilesional+motor+cortical+excitability+post-stroke.&aulast=Li. doi:10.1080/10749357.2019.1623458.

Lin JH HI, Sheu CF, Hsieh CL. 2004. Psychometric properties of the sensory scale of the fugl-meyer assessment in stroke patients. Clinical Rehabilitation 18(391 397.

Lin K-C, Chuang L-L, Wu C-Y, Hsieh Y-W, Chang W-Y. 2010. Responsiveness and validity of three dexterous function measures in stroke rehabilitation. The Journal of Rehabilitation Research and Development. 47(6):563. https://dx.doi.org/10.1682/jrrd.2009.09.0155. doi:10.1682/jrrd.2009.09.0155.

Lulic T, El-Sayes J, Fassett HJ, Nelson AJ. 2017. Physical activity levels determine exercise-induced changes in brain excitability. PLoS ONE [Electronic Resource]. 12(3):e0173672. https://proxy.library.mcgill.ca/login?url=http%3A%2F%2Fovidsp.ovid.com/ovidweb.cgi?T=JS&CSC=Y&NEWS=N&PAGE=fulltext&D=med14&AN=28278300 http://mcgill.on.worldcat.org/atoztitles/link?sid=OVID:medline&id=pmid:28278300&id=doi:10.1371%2Fjournal.pone.0173672&issn=1932-6203&isbn=&volume=12&issue=3&spage=e0173672&pages=e0173672&date=2017&title=PLoS+ONE+%5BElectronic+Resource%5D&atitle=Physical+activity+levels+determine+exercise-induced+changes+in+brain+excitability.&aulast=Lulic. doi:10.1371/journal.pone.0173672.

Luo L et al. 2020. Effect of high-intensity exercise on cardiorespiratory fitness in stroke survivors: A systematic review and meta-analysis. Ann Phys Rehabil Med. 63(1):59–68. https://www.ncbi.nlm.nih.gov/pubmed/31465865. doi:10.1016/j.rehab.2019.07.006.

Lyden PD, Hedges B. 1992. Protective effect of synaptic inhibition during cerebral ischemia in rats and rabbits. Stroke. 23(10):1463–1469. https://dx.doi.org/10.1161/01.str.23.10.1463. doi:10.1161/01.str.23.10.1463.

Maldonado MA, Allred RP, Felthauser EL, Jones TA. 2008. Motor skill training, but not voluntary exercise, improves skilled reaching after unilateral ischemic lesions of the sensorimotor cortex in rats. Neurorehabilitation and Neural Repair. 22(3):250–261. https://dx.doi.org/10.1177/1545968307308551. doi:10.1177/1545968307308551.

Mang CS, Snow NJ, Campbell KL, Ross CJD, Boyd LA. 2014. A single bout of high-intensity aerobic exercise facilitates response to paired associative stimulation and promotes sequence-specific implicit motor learning. J Appl Physiol. 117(11):1325–1336. https://dx.doi.org/10.1152/japplphysiol.00498.2014. doi:10.1152/japplphysiol.00498.2014.

Marin R et al. 2003. The effect of voluntary exercise exposure on histological and neurobehavioral outcomes after ischemic brain injury in the rat. Physiol Behav. 80(2-3):167–175. https://www.ncbi.nlm.nih.gov/pubmed/14637213. doi:10.1016/j.physbeh.2003.06.001.

Marzolini S. 2021. Associations between time after stroke and exercise training outcomes: A metaregression analysis. Journal of the American Heart Association. doi:10.1161/JAHA.121.022588.

McDonnell MN, Stinear CM. 2017. Tms measures of motor cortex function after stroke: A meta-analysis. Brain Stimul. 10(4):721–734. https://dx.doi.org/10.1016/j.brs.2017.03.008. doi:10.1016/j.brs.2017.03.008.

Moncion K et al. 2024. Cardiorespiratory fitness benefits of high-intensity interval training after stroke: A randomized controlled trial. Stroke. https://dx.doi.org/10.1161/strokeaha.124.046564. doi:10.1161/strokeaha.124.046564.

Morris TP et al. 2020. Light aerobic exercise modulates executive function and cortical excitability. Eur J Neurosci. 51(7):1723–1734. https://proxy.library.mcgill.ca/login?url=http%3A%2F%2Fovidsp.ovid.com/ovidweb.cgi?T=JS&CSC=Y&NEWS=N&PAGE=fulltext&D=medl&AN=31605625 http://mcgill.on.worldcat.org/atoztitles/link?sid=OVID:medline&id=pmid:31605625&id=doi:10.1111%2Fejn.14593&issn=0953-816X&isbn=&volume=51&issue=7&spage=1723&pages=1723-1734&date=2020&title=European+Journal+of+Neuroscience&atitle=Light+aerobic+exercise+modulates+executive+function+and+cortical+excitability.&aulast=Morris. doi:10.1111/ejn.14593.

Murphy TH, Corbett D. 2009. Plasticity during stroke recovery: From synapse to behaviour. Nat Rev Neurosci. 10(12):861–872. https://dx.doi.org/10.1038/nrn2735. 10.1038/nrn2735.

Nepveu J-F et al. 2017. A single bout of high-intensity interval training improves motor skill retention in individuals with stroke. Neurorehabilitation and Neural Repair. 31(8):726–735. https://dx.doi.org/10.1177/1545968317718269. doi:10.1177/1545968317718269.

Neva JL, Brown KE, Mang CS, Francisco BA, Boyd LA. 2017. An acute bout of exercise modulates both intracortical and interhemispheric excitability. Eur J Neurosci. 45(10):1343–1355. https://proxy.library.mcgill.ca/login?url=http%3A%2F%2Fovidsp.ovid.com/ovidweb.cgi?T=JS&CSC=Y&NEWS=N&PAGE=fulltext&D=med14&AN=28370664 http://mcgill.on.worldcat.org/atoztitles/link?sid=OVID:medline&id=pmid:28370664&id=doi:10.1111%2Fejn.13569&issn=0953-816X&isbn=&volume=45&issue=10&spage=1343&pages=1343-1355&date=2017&title=European+Journal+of+Neuroscience&atitle=An+acute+bout+of+exercise+modulates+both+intracortical+and+interhemispheric+excitability.&aulast=Neva. doi:10.1111/ejn.13569.

Nicolini C et al. 2019. No changes in corticospinal excitability, biochemical markers, and working memory after six weeks of high-intensity interval training in sedentary males. Physiological Reports. 7(11):e14140. https://proxy.library.mcgill.ca/login?url=http%3A%2F%2Fovidsp.ovid.com/ovidweb.cgi?T=JS&CSC=Y&NEWS=N&PAGE=fulltext&D=med16&AN=31175708 http://mcgill.on.worldcat.org/atoztitles/link?sid=OVID:medline&id=pmid:31175708&id=doi:10.14814%2Fphy2.14140&issn=2051-817X&isbn=&volume=7&issue=11&spage=e14140&pages=e14140&date=2019&title=Physiological+Reports&atitle=No+changes+in+corticospinal+excitability%2C+biochemical+markers%2C+and+working+memory+after+six+weeks+of+high-intensity+interval+training+in+sedentary+males.&aulast=Nicolini. doi:10.14814/phy2.14140.

Orfila JE et al. 2019. Delayed inhibition of tonic inhibition enhances functional recovery following experimental ischemic stroke. Journal of Cerebral Blood Flow & Metabolism. 39(6):1005–1014. https://dx.doi.org/10.1177/0271678x17750761. doi:10.1177/0271678x17750761.

Ostadan F et al. 2016. Changes in corticospinal excitability during consolidation predict acute exercise-induced off-line gains in procedural memory. Neurobiology of Learning and Memory. 136(196–203. https://dx.doi.org/10.1016/j.nlm.2016.10.009. doi:10.1016/j.nlm.2016.10.009.

Perez MA, Lundbye-Jensen J, Nielsen JB. 2006. Changes in corticospinal drive to spinal motoneurones following visuo-motor skill learning in humans. J Physiol. 573(Pt 3):843–855. https://www.ncbi.nlm.nih.gov/pubmed/16581867. doi:10.1113/jphysiol.2006.105361.

Plautz EJ, Milliken GW, Nudo RJ. 2000. Effects of repetitive motor training on movement representations in adult squirrel monkeys: Role of use versus learning. Neurobiology of Learning and Memory. 74(1):27–55. https://dx.doi.org/10.1006/nlme.1999.3934. doi:10.1006/nlme.1999.3934.

Ploughman M, Attwood Z, White N, Dore JJ, Corbett D. 2007. Endurance exercise facilitates relearning of forelimb motor skill after focal ischemia. Eur J Neurosci. 25(11):3453–3460. https://www.ncbi.nlm.nih.gov/pubmed/17553014. doi:10.1111/j.1460-9568.2007.05591.x.

Ploughman M, Austin MW, Glynn L, Corbett D. 2015. The effects of poststroke aerobic exercise on neuroplasticity: A systematic review of animal and clinical studies. Transl Stroke Res. 6(1):13–28. doi:10.1007/s12975-014-0357-7.

Ploughman M, Kelly LP. 2016. Four birds with one stone? Reparative, neuroplastic, cardiorespiratory, and metabolic benefits of aerobic exercise poststroke. Curr Opin Neurol. 29(6):684–692. https://www.ncbi.nlm.nih.gov/pubmed/27661010. doi:10.1097/WCO.0000000000000383.

Robertson EM, Takacs A. 2017. Exercising control over memory consolidation. Trends Cogn Sci. 21(5):310–312. https://dx.doi.org/10.1016/j.tics.2017.03.001. doi:10.1016/j.tics.2017.03.001.

Rodrigues L et al. 2025. Modulating brain excitability with cardiovascular exercise in chronic stroke: A randomized controlled trial. Neurorehabil Neural Repair.15459683251351883. https://www.ncbi.nlm.nih.gov/pubmed/40637167. doi:10.1177/15459683251351883.

Rossini PM et al. 2015. Non-invasive electrical and magnetic stimulation of the brain, spinal cord, roots and peripheral nerves: Basic principles and procedures for routine clinical and research application. An updated report from an i.F.C.N. Committee. Clin Neurophysiol. 126(6):1071–1107. https://dx.doi.org/10.1016/j.clinph.2015.02.001. doi:10.1016/j.clinph.2015.02.001.

Singh AM, Duncan RE, Neva JL, Staines W. 2014. Aerobic exercise modulates intracortical inhibition and facilitation in a nonexercised upper limb muscle. 6(1):23. https://dx.doi.org/10.1186/2052-1847-6-23. doi:10.1186/2052-1847-6-23.

Singh AM, Staines WR. 2015. The effects of acute aerobic exercise on the primary motor cortex. J Mot Behav. 47(4):328–339. https://www.ncbi.nlm.nih.gov/pubmed/25565153. doi:10.1080/00222895.2014.983450.

Sivaramakrishnan A, Subramanian SK. 2023. A systematic review on the effects of acute aerobic exercise on neurophysiological, molecular, and behavioral measures in chronic stroke. Neurorehabil Neural Repair. 37(2-3):151–164. https://www.ncbi.nlm.nih.gov/pubmed/36703562. doi:10.1177/15459683221146996.

Smith AE, Goldsworthy MR, Garside T, Wood FM, Ridding MC. 2014. The influence of a single bout of aerobic exercise on short-interval intracortical excitability. Exp Brain Res. 232(6):1875–1882. https://proxy.library.mcgill.ca/login?url=http%3A%2F%2Fovidsp.ovid.com/ovidweb.cgi?T=JS&CSC=Y&NEWS=N&PAGE=fulltext&D=med11&AN=24570388 http://mcgill.on.worldcat.org/atoztitles/link?sid=OVID:medline&id=pmid:24570388&id=doi:10.1007%2Fs00221-014-3879-z&issn=0014-4819&isbn=&volume=232&issue=6&spage=1875&pages=1875-82&date=2014&title=Experimental+Brain+Research&atitle=The+influence+of+a+single+bout+of+aerobic+exercise+on+short-interval+intracortical+excitability.&aulast=Smith. doi:10.1007/s00221-014-3879-z.

Stagg CJ et al. 2011. Relationship between physiological measures of excitability and levels of glutamate and gaba in the human motor cortex. The Journal of Physiology. 589(23):5845–5855. https://dx.doi.org/10.1113/jphysiol.2011.216978. doi:10.1113/jphysiol.2011.216978.

Stavrinos EL, Coxon JP. 2017. High-intensity interval exercise promotes motor cortex disinhibition and early motor skill consolidation. Journal of Cognitive Neuroscience. 29(4):593–604. https://proxy.library.mcgill.ca/login?url=http%3A%2F%2Fovidsp.ovid.com/ovidweb.cgi?T=JS&CSC=Y&NEWS=N&PAGE=fulltext&D=med14&AN=27897671 http://mcgill.on.worldcat.org/atoztitles/link?sid=OVID:medline&id=pmid:27897671&id=doi:10.1162%2Fjocn_a_01078&issn=0898-929X&isbn=&volume=29&issue=4&spage=593&pages=593-604&date=2017&title=Journal+of+Cognitive+Neuroscience&atitle=High-intensity+Interval+Exercise+Promotes+Motor+Cortex+Disinhibition+and+Early+Motor+Skill+Consolidation.&aulast=Stavrinos. doi:10.1162/jocn_a_01078.

Stinear CM et al. 2006. Functional potential in chronic stroke patients depends on corticospinal tract integrity. Brain. 130(1):170–180. https://dx.doi.org/10.1093/brain/awl333. doi:10.1093/brain/awl333.

Stinear CM et al. 2017. Prep2: A biomarker-based algorithm for predicting upper limb function after stroke. Annals of Clinical and Translational Neurology. 4(11):811–820. https://dx.doi.org/10.1002/acn3.488. doi:10.1002/acn3.488.

Szuhany KL, Bugatti M, Otto MW. 2015. A meta-analytic review of the effects of exercise on brain-derived neurotrophic factor. J Psychiatr Res. 60(56–64. https://dx.doi.org/10.1016/j.jpsychires.2014.10.003. doi:10.1016/j.jpsychires.2014.10.003.

Talelli P, Greenwood RJ, Rothwell JC. 2006. Arm function after stroke: Neurophysiological correlates and recovery mechanisms assessed by transcranial magnetic stimulation. 117(8):1641–1659. https://dx.doi.org/10.1016/j.clinph.2006.01.016. doi:10.1016/j.clinph.2006.01.016.

Thickbroom GW et al. 2015. Stroke subtype and motor impairment influence contralesional excitability. Neurology. 85(6):517–520. https://dx.doi.org/10.1212/wnl.0000000000001828. doi:10.1212/wnl.0000000000001828.

Tunovic S, Press DZ, Robertson EM. 2014. A physiological signal that prevents motor skill improvements during consolidation. J Neurosci. 34(15):5302–5310. https://dx.doi.org/10.1523/JNEUROSCI.3497-13.2014. doi:10.1523/jneurosci.3497-13.2014.

Voss MW, Vivar C, Kramer AF, Van Praag H. 2013. Bridging animal and human models of exercise-induced brain plasticity. Trends Cogn Sci. 17(10):525–544. https://dx.doi.org/10.1016/j.tics.2013.08.001. doi:10.1016/j.tics.2013.08.001.

Wang Y-C et al. 2018. Postacute delivery of gaba a α5 antagonist promotes postischemic neurological recovery and peri-infarct brain remodeling. Stroke. 49(10):2495–2503. https://dx.doi.org/10.1161/strokeaha.118.021378. doi:10.1161/strokeaha.118.021378.

Winstein C, Lewthwaite R, Blanton SR, Wolf LB, Wishart L. 2014. Infusing motor learning research into neurorehabilitation practice: A historical perspective with case exemplar from the accelerated skill acquisition program. J Neurol Phys Ther. 38(3):190–200. <Go to ISI>://WOS:000339053700008. doi:10.1097/npt.0000000000000046.

Yamazaki Y et al. 2019. Acute low-intensity aerobic exercise modulates intracortical inhibitory and excitatory circuits in an exercised and a non-exercised muscle in the primary motor cortex. Front Physiol. 10(https://dx.doi.org/10.3389/fphys.2019.01361. doi:10.3389/fphys.2019.01361.

Yang Y-R, Chen IH, Liao K-K, Huang C-C, Wang R-Y. 2010. Cortical reorganization induced by body weight–supported treadmill training in patients with hemiparesis of different stroke durations. Arch Phys Med Rehabil. 91(4):513–518. https://dx.doi.org/10.1016/j.apmr.2009.11.021. doi:10.1016/j.apmr.2009.11.021.

Yang YR, Wang RY, Wang PS. 2003. Early and late treadmill training after focal brain ischemia in rats. Neurosci Lett. 339(2):91–94. https://www.ncbi.nlm.nih.gov/pubmed/12614902. doi:10.1016/s0304-3940(03)00010-7.

Yen C-L, Wang R-Y, Liao K-K, Huang C-C, Yang Y-R. 2008. Gait training-induced change in corticomotor excitability in patients with chronic stroke. Neurorehabilitation and Neural Repair. 22(1):22–30. <Go to ISI>://WOS:000251732300002. doi:10.1177/1545968307301875.

Ziemann U et al. 2015. Tms and drugs revisited 2014. Clin Neurophysiol. 126(10):1847–1868. https://www.ncbi.nlm.nih.gov/pubmed/25534482. doi:10.1016/j.clinph.2014.08.028.

