## Supplementary material for "The Effects of Cardiovascular Exercise on Corticospinal Excitability in People with Subacute Stroke": Suplementary Materials

**Supplementary Table 1.** Estimated within-group changes in cardiorespiratory fitness, upper-limb impairment and function from T0 to T2.

| | T0 | T1 | T2 |
|---|---|---|---|
| **GXT ($VO_2$peak, mL.Kg$^{-1}$.min$^{-1}$)** | | | |
| **CE+Standard care** | 17.05 (0.84) | 19.86 (0.85) | 21.52 (0.86) |
| **Standard care** | 17.80 (0.99) | 17.93 (1.01) | 18.17 (1.03) |
| **UL-FMA** | | | |
| **CE+Standard care** | 56.54 (1.14) | 59.07 (1.16) | 60.29 (1.17) |
| **Standard care** | 59.06 (1.35) | 60.88 (1.37) | 60.87 (1.40) |
| **BBT** | | | |
| **CE+Standard care** | 45.81 (1.93) | 48.51 (1.95) | 50.38 (1.97) |
| **Standard care** | 48.22 (2.26) | 51.77 (2.31) | 53.06 (2.35) |

BBT, box and block Test; CE, cardiovascular exercise; UL-FMA, upper-limb Fugl-Meyer assessment; GXT, graded exercise test; mL.Kg$^{-1}$.min$^{-1}$, milliliters per kilogram per minute. Data are presented as least squares means with standard errors (SE).

**Supplementary Table 2.** Estimated within-group chronic changes and between-group differences in corticospinal excitability measures from T0 to T2.

| | CE+standard care | | Standard care | | Between-Group Differences | |
|---|---|---|---|---|---|---|
| **CSE Outcome** | **Estimate (95% CI)** | **p value** | **Estimate (95% CI)** | **p value** | **Estimate (95% CI)** | **p value** |
| **RMT** | | | | | | |
| **Ipsilesional (%)** | -0.44 (-3.22 to 2.22) | 0.66 | -0.24 (-3.73 to 3.24) | 0.24 | -0.20 (−4.62 to 4.22) | 0.19 |
| **Contralesional (%)** | 0.30 (-1.17 to 1.77) | 0.43 | 0.75 (-2.53 to 4.03) | 0.83 | -0.45 (−3.86 to 2.96) | 0.70 |
| **MEP amplitude (resting)** | | | | | | |
| **Ipsilesional (mV)** | -0.10 (-0.28 to 0.07) | 0.31 | -0.009 (-0.11 to 0.09) | 0.80 | -0.091 (−0.29 to 0.11) | 0.55 |
| **Contralesional (mV)** | -0.003 (-0.10 to 0.09) | 0.99 | -0.09 (-0.23 to 0.04) | 0.24 | 0.087 (−0.07 to 0.25) | 0.45 |
| **MEP amplitude (active)** | | | | | | |
| **Ipsilesional (mV)** | -0.22 (-0.47 to 0.01) | 0.06 | -0.07 (-0.11 to 0.09) | 0.33 | −0.15 (−0.41 to 0.11) | 0.66 |
| **Contralesional (mV)** | -0.03 (-0.38 to 0.30) | 0.94 | -0.25 (-0.60 to 0.08) | 0.10 | 0.22 (−0.26 to 0.70) | 0.40 |
| **CSP** | | | | | | |
| **Ipsilesional (ms)** | -0.004 (-0.01 to 0.007) | 0.60 | -0.003 (-0.01 to 0.007) | 0.27 | −0.001 (−0.01 to 0.01) | 0.54 |
| **Contralesional (ms)** | 0.001 (-0.005 to 0.008) | 0.25 | -0.005 (-0.01 to 0.005) | 0.48 | 0.006 (−0.003 to 0.01) | 0.18 |
| **ICF** | | | | | | |
| **Ipsilesional** | 0.69 (-0.57 to 1.96) | 0.36 | 0.18 (-0.79 to 1.15) | 0.88 | 0.51 (−1.08 to 2.10) | 0.74 |
| **Contralesional** | -0.60 (-1.59 to 0.39) | 0.34 | 0.44 (-1.02 to 1.90) | 0.47 | −1.04 (−2.80 to 0.72) | 0.19 |
| **SICI** | | | | | | |
| **Ipsilesional** | 0.005 (-0.36 to 0.37) | 0.97 | 0.12 (-0.37 to 0.62) | 0.24 | −0.11 (0.73 to 0.5) | 0.32 |
| **Contralesional** | -0.39 (-1.12 to 0.34) | 0.32 | 0.02 (-0.55 to 0.61) | 0.96 | −0.41 (−1.34 to 0.52) | 0.54 |

CE, cardiovascular exercise; CI, confidence interval; CSE, corticospinal excitability; CSP, cortical silent period; ICF, intracortical facilitation; MEP, motor evoked potential; ms, millisecond; mV, millivolt; RMT, resting motor threshold; SICI, short-intracortical inhibition. Data is presented as least squares means with 95% confidence intervals.

**Supplementary Table 3.** Estimated within-group acute changes and between-group differences in corticospinal excitability measures from T0 to T2.

| | CE+standard care | | Standard care | | Between-Group Differences | |
|---|---|---|---|---|---|---|
| **CSE Outcome** | **Estimate (95% CI)** | **p value** | **Estimate (95% CI)** | **p value** | **Estimate (95% CI)** | **p value** |
| **MEP amplitude (resting)** | | | | | | |
| **Ipsilesional (mV)** | 0.03 (-0.13 to 0.21) | 0.86 | -0.006 (-0.17 to 0.16) | 0.59 | 0.03 (−0.20 to 0.27) | 0.69 |
| **Contralesional (mV)** | -0.08 (-0.23 to 0.05) | 0.34 | -0.01 (-0.19 to 0.15) | 0.38 | −0.07 (−0.29 to 0.15) | 0.34 |
| **MEP amplitude (active)** | | | | | | |
| **Ipsilesional (mV)** | 0.16 (-0.02 to 0.35) | 0.070 | 0.02 (-0.17 to 0.21) | 0.96 | 0.14 (−0.12 to 0.40) | 0.13 |
| **Contralesional (mV)** | -0.26 (-0.53 to 0.002) | 0.052 | 0.006 (-0.25 to 0.26) | 0.60 | −0.26 (−0.63 to 0.10) | 0.23 |
| **CSP** | | | | | | |
| **Ipsilesional (ms)** | -0.003 (-0.02 to 0.02) | 0.82 | 0.003 (-0.007 to 0.01) | 0.37 | −0.006 (−0.02 to 0.01) | 0.75 |
| **Contralesional (ms)** | -0.0026 (-0.01 to 0.006) | 0.68 | -0.0023 (-0.01 to 0.009) | 0.67 | -0.0003 (−0.01 to 0.01) | 0.99 |
| **ICF** | | | | | | |
| **Ipsilesional** | 0.02 (-1.21 to 1.26) | 0.83 | -0.18 (-1.76 to 1.38) | 0.95 | 0.20 (−2.13 to 2.53) | 0.87 |
| **Contralesional** | 0.34 (-0.59 to 1.28) | 0.66 | 0.14 (-1.38 to 1.66) | 0.88 | 0.20 (−1.58 to 1.98) | 0.84 |
| **SICI** | | | | | | |
| **Ipsilesional** | 0.04 (-0.48 to 0.57) | 0.97 | -0.14 (-0.70 to 0.41) | 0.67 | 0.18 (−0.58 to 0.94) | 0.69 |
| **Contralesional** | 0.30 (-0.32 to 0.94) | 0.39 | 0.11 (-0.43 to 0.67) | 0.66 | 0.19 (−0.64 to 1.02) | 0.92 |

CE, cardiovascular exercise; CI, confidence interval; CSE, corticospinal excitability; CSP, cortical silent period; ICF, intracortical facilitation; MEP, motor evoked potential; ms, millisecond; mV, millivolt; SICI, short-intracortical inhibition. Data is presented as least squares means with 95% confidence intervals.

**Supplementary Table 4.** Estimated within-group acute changes in corticospinal excitability measures at T0 (n=74).

| CSE Outcome | Estimate (95% CI) | p value |
|---|---|---|
| **MEP amplutude (resting)** | | |
| **Ipsilesional (mV)** | 0.002 (-0.03 to 0.04) | 0.902 |
| **Contralesional (mV)** | 0.04 (0.01 to 0.08) | 0.006* |
| **MEP amplutude (active)** | | |
| **Ipsilesional (mV)** | -0.01 (-0.06 to 0.02) | 0.340 |
| **Contralesional (mV)** | 0.06 (0.01 to 0.11) | 0.008* |
| **CSP** | | |
| **Ipsilesional (ms)** | 2.614e-5 (-0.003 to 0.003) | 0.987 |
| **Contralesional (ms)** | 0.0003 (-0.001 to 0.002) | 0.687 |
| **ICF** | | |
| **Ipsilesional** | -0.003 (-0.18 to 0.18) | 0.971 |
| **Contralesional** | -0.03 (-0.29 to 0.22) | 0.772 |
| **SICI** | | |
| **Ipsilesional** | 0.04 (-0.06 to 0.14) | 0.413 |
| **Contralesional** | 0.02 (-0.04 to 0.10) | 0.461 |

CI, confidence interval; CSE, corticospinal excitability; CSP, cortical silent period; ICF, intracortical facilitation; MEP, motor evoked potential; ms, millisecond; mV, millivolt; SICI, short-intracortical inhibition. Data is presented as least squares means with 95% confidence intervals. * $p<0.05$

**Supplementary Table 5.** Adjusted linear mixed models comparing chronic corticospinal excitability responses to cardiovascular exercise training across different lesion location groups—cortical, subcortical, and cerebellar—in both ipsilesional and contralesional hemispheres.

| | DFNum | DFDen | F ratio | p value |
|---|---|---|---|---|
| **RMT ipsilesional** | | | | |
| **Group** | 1 | 63.5 | 1e-5 | **0.997** |
| **Time** | 2 | 104.2 | 0.50 | **0.605** |
| **Time*Group[Location]** | 6 | 104.4 | 0.57 | **0.747** |
| **Age** | 1 | 62.6 | 0.01 | **0.920** |
| **Sex** | 1 | 62.6 | 1.24 | **0.269** |
| **NIHSS** | 1 | 63.3 | 4.79 | **0.032*** |
| **MVC** | 1 | 62.2 | 5.42 | **0.023*** |
| **Resting MEP ipsilesional** | | | | |
| **Group** | 1 | 63.5 | 2.48 | **0.120** |
| **Time** | 2 | 81.5 | 0.40 | **0.669** |
| **Time*Group[Location]** | 6 | 88.6 | 0.64 | **0.698** |
| **Age** | 1 | 59.9 | 0.41 | **0.520** |
| **Sex** | 1 | 59.0 | 0.32 | **0.573** |
| **NIHSS** | 1 | 60.9 | 1.002 | **0.321** |
| **MVC** | 1 | 57.8 | 10.31 | **0.002*** |
| **Active MEP ipsilesional** | | | | |
| **Group** | 1 | 64.2 | 2.09 | **0.152** |
| **Time** | 2 | 102.5 | 2.89 | **0.060** |
| **Time*Group[Location]** | 6 | 102.4 | 0.75 | **0.611** |
| **Age** | 1 | 62.1 | 0.70 | **0.406** |
| **Sex** | 1 | 61.4 | 0.013 | **0.909** |
| **NIHSS** | 1 | 62.9 | 0.64 | **0.427** |
| **MVC** | 1 | 60.7 | 16.02 | **0.003*** |
| **CSP ipsilesional** | | | | |
| **Group** | 1 | 64.3 | 0.13 | **0.715** |
| **Time** | 2 | 101.4 | 0.79 | **0.455** |
| **Time*Group[Location]** | 6 | 101.4 | 1.09 | **0.371** |
| **Age** | 1 | 62.7 | 0.55 | **0.459** |
| **Sex** | 1 | 62.3 | 7.38 | **0.009*** |
| **NIHSS** | 1 | 63.3 | 0.67 | **0.416** |
| **MVC** | 1 | 61.8 | 3.09 | **0.083** |
| **ICF ipsilesional** | | | | |
| **Group** | 1 | 66.3 | 0.90 | **0.346** |
| **Time** | 2 | 86.9 | 0.40 | **0.671** |
| **Time*Group[Location]** | 6 | 94.8 | 1.67 | **0.137** |
| **Age** | 1 | 64.0 | 0.05 | **0.812** |
| **Sex** | 1 | 62.5 | 0.002 | **0.958** |
| **NIHSS** | 1 | 64.8 | 2.68 | **0.106** |
| **MVC** | 1 | 60.9 | 0.20 | **0.651** |
| **SICI ipsilesional** | | | | |
| **Group** | 1 | 54.7 | 0.71 | **0.401** |

| | | | | |
|---|---|---|---|---|
| **Time** | 2 | 69.2 | 0.71 | **0.492** |
| **Time*Group[Location]** | 6 | 76.1 | 0.67 | **0.674** |
| **Age** | 1 | 52.4 | 0.77 | **0.382** |
| **Sex** | 1 | 52.8 | 1.24 | **0.270** |
| **NIHSS** | 1 | 54.6 | 0.77 | **0.382** |
| **MVC** | 1 | 52.9 | 3.56 | **0.065** |
| **RMT contralesional** | | | | |
| **Group** | 1 | 67.8 | 0.02 | **0.872** |
| **Time** | 2 | 111.3 | 0.24 | **0.781** |
| **Time*Group[Location]** | 6 | 111.6 | 0.50 | **0.805** |
| **Age** | 1 | 66.6 | 0.19 | **0.661** |
| **Sex** | 1 | 67.0 | 4.78 | **0.032*** |
| **NIHSS** | 1 | 67.5 | 0.68 | **0.413** |
| **MVC** | 1 | 66.5 | 0.003 | **0.957** |
| **Resting MEP contralesional** | | | | |
| **Group** | 1 | 67.4 | 0.32 | **0.573** |
| **Time** | 2 | 112.3 | 1.04 | **0.356** |
| **Time*Group[Location]** | 6 | 112.7 | 0.90 | **0.491** |
| **Age** | 1 | 64.2 | 0.001 | **0.979** |
| **Sex** | 1 | 65.7 | 0.22 | **0.641** |
| **NIHSS** | 1 | 66.7 | 0.02 | **0.869** |
| **MVC** | 1 | 63.9 | 2.26 | **0.137** |
| **Active MEP contralesional** | | | | |
| **Group** | 1 | 63.0 | 1.19 | **0.279** |
| **Time** | 2 | 80.5 | 0.73 | **0.481** |
| **Time*Group[Location]** | 6 | 92.2 | 0.97 | **0.446** |
| **Age** | 1 | 57.8 | 0.08 | **0.769** |
| **Sex** | 1 | 60.0 | 0.41 | **0.520** |
| **NIHSS** | 1 | 61.3 | 0.83 | **0.364** |
| **MVC** | 1 | 57.5 | 1.62 | **0.207** |
| **CSP contralesional** | | | | |
| **Group** | 1 | 67.2 | 0.37 | **0.543** |
| **Time** | 2 | 112.1 | 0.54 | **0.579** |
| **Time*Group[Location]** | 6 | 112.5 | 0.73 | **0.626** |
| **Age** | 1 | 64.1 | 0.62 | **0.431** |
| **Sex** | 1 | 65.6 | 3.33 | **0.072** |
| **NIHSS** | 1 | 66.5 | 0.32 | **0.574** |
| **MVC** | 1 | 63.8 | 0.03 | **0.843** |
| **ICF contralesional** | | | | |
| **Group** | 1 | 71.3 | 1.44 | **0.234** |
| **Time** | 2 | 96.0 | 0.33 | **0.719** |
| **Time*Group[Location]** | 6 | 107.0 | 0.44 | **0.849** |
| **Age** | 1 | 65.1 | 1.08 | **0.301** |
| **Sex** | 1 | 67.7 | 0.75 | **0.389** |
| **NIHSS** | 1 | 70.1 | 0.02 | **0.879** |
| **MVC** | 1 | 64.1 | 0.23 | **0.633** |
| **SICI contralesional** | | | | |
| **Group** | 1 | 57.3 | 1.39 | **0.242** |
| **Time** | 2 | 70.1 | 0.06 | **0.940** |

| | | | | |
|---|---|---|---|---|
| **Time*Group[Location]** | 6 | 82.7 | 0.22 | **0.966** |
| **Age** | 1 | 54.2 | 0.08 | **0.776** |
| **Sex** | 1 | 55.9 | 0.01 | **0.903** |
| **NIHSS** | 1 | 58.6 | 0.82 | **0.366** |
| **MVC** | **1** | **54.7** | **2.51** | **0.118** |

CSP, cortical silent period; ICF, intracortical facilitation; MEP, motor evoked potential; MVC, maximal voluntary contraction; NIHSS, national institutes of health stroke scale; RMT, resting motor threshold; SICI, short-intracortical inhibition. Location represents the three lesion location groups—cortical, subcortical, and cerebellar—nested in the model as a categorical variable. * $p<0.05$

**Supplementary Table 6.** Adjusted linear mixed models comparing acute corticospinal excitability responses to cardiovascular exercise training across different lesion location groups—cortical, subcortical, and cerebellar—in both ipsilesional and contralesional hemispheres.

| | DFNum | DFDen | F ratio | p value |
|---|---|---|---|---|
| **Resting MEP ipsilesional** | | | | |
| **Group** | 1 | 63.4 | 1.76 | **0.189** |
| **Time** | 2 | 85.7 | 0.16 | **0.849** |
| **Time*Group[Location]** | 6 | 93.4 | 0.43 | **0.851** |
| **Age** | 1 | 62.3 | 2e-4 | **0.989** |
| **Sex** | 1 | 60.6 | 0.25 | **0.615** |
| **NIHSS** | 1 | 63.3 | 0.81 | **0.371** |
| **MVC** | 1 | 59.2 | 0.04 | **0.831** |
| **Active MEP ipsilesional** | | | | |
| **Group** | 1 | 54.9 | 4.50 | **0.038*** |
| **Time** | 2 | 80.5 | 4.66 | **0.012*** |
| **Time*Group[Location]** | 6 | 85.5 | 0.83 | **0.543** |
| **Age** | 1 | 52.1 | 0.35 | **0.552** |
| **Sex** | 1 | 50.9 | 0.35 | **0.556** |
| **NIHSS** | 1 | 53.0 | 1.49 | **0.227** |
| **MVC** | 1 | 49.9 | 0.54 | **0.462** |
| **CSP ipsilesional** | | | | |
| **Group** | 1 | 50.5 | 0.73 | **0.394** |
| **Time** | 2 | 106.6 | 0.62 | **0.539** |
| **Time*Group[Location]** | 6 | 106.6 | 1.13 | **0.350** |
| **Age** | 1 | 50.2 | 3.17 | **0.081** |
| **Sex** | 1 | 48.7 | 1.64 | **0.206** |
| **NIHSS** | 1 | 51.0 | 1.52 | **0.222** |
| **MVC** | 1 | 48.5 | 0.45 | **0.505** |
| **ICF ipsilesional** | | | | |
| **Group** | 1 | 62.4 | 0.01 | **0.913** |
| **Time** | 2 | 111.1 | 0.06 | **0.934** |
| **Time*Group[Location]** | 6 | 110.1 | 1.64 | **0.142** |
| **Age** | 1 | 60.7 | 0.17 | **0.677** |
| **Sex** | 1 | 59.0 | 5e-4 | **0.983** |
| **NIHSS** | 1 | 61.6 | 0.006 | **0.937** |
| **MVC** | 1 | 57.5 | 0.47 | **0.495** |
| **SICI ipsilesional** | | | | |
| **Group** | 1 | 54.5 | 3.11 | **0.083** |
| **Time** | 2 | 73.6 | 0.07 | **0.924** |
| **Time*Group[Location]** | 6 | 80.5 | 0.61 | **0.722** |
| **Age** | 1 | 52.8 | 0.75 | **0.388** |
| **Sex** | 1 | 53.1 | 2.84 | **0.098** |
| **NIHSS** | 1 | 55.3 | 2.55 | **0.115** |
| **MVC** | 1 | 53.0 | 1.27 | **0.264** |
| **Resting MEP contralesional** | | | | |
| **Group** | 1 | 70.4 | 0.22 | **0.634** |

| | | | | |
|---|---|---|---|---|
| **Time** | 2 | 122.8 | 0.65 | **0.520** |
| **Time*Group[Location]** | 6 | 123.7 | 0.60 | **0.724** |
| **Age** | 1 | 65.2 | 0.02 | **0.880** |
| **Sex** | 1 | 67.1 | 0.005 | **0.943** |
| **NIHSS** | 1 | 69.7 | 0.71 | **0.400** |
| **MVC** | 1 | 64.2 | 0.19 | **0.663** |
| **Active MEP contralesional** | | | | |
| **Group** | 1 | 58.6 | 0.03 | **0.856** |
| **Time** | 2 | 96.8 | 2.49 | **0.088** |
| **Time*Group[Location]** | 6 | 104.5 | 1.82 | **0.102** |
| **Age** | 1 | 56.2 | 3.57 | **0.064** |
| **Sex** | 1 | 57.1 | 0.02 | **0.869** |
| **NIHSS** | 1 | 59.6 | 0.17 | **0.673** |
| **MVC** | 1 | 55.3 | 1.61 | **0.209** |
| **CSP contralesional** | | | | |
| **Group** | 1 | 62.3 | 1.37 | **0.246** |
| **Time** | 2 | 122.1 | 0.19 | **0.825** |
| **Time*Group[Location]** | 6 | 123.3 | 0.67 | **0.671** |
| **Age** | 1 | 58.0 | 2.65 | **0.109** |
| **Sex** | 1 | 59.2 | 0.70 | **0.403** |
| **NIHSS** | 1 | 61.8 | 0.31 | **0.574** |
| **MVC** | 1 | 57.2 | 0.01 | **0.910** |
| **ICF contralesional** | | | | |
| **Group** | 1 | 65.2 | 0.67 | **0.413** |
| **Time** | 2 | 93.9 | 0.16 | **0.850** |
| **Time*Group[Location]** | 6 | 105.5 | 0.13 | **0.991** |
| **Age** | 1 | 61.1 | 0.48 | **0.488** |
| **Sex** | 1 | 62.4 | 0.01 | **0.901** |
| **NIHSS** | 1 | 65.2 | 1e-4 | **0.991** |
| **MVC** | 1 | 59.7 | 0.03 | **0.844** |
| **SICI contralesional** | | | | |
| **Group** | 1 | 53.7 | 0.17 | **0.677** |
| **Time** | 2 | 80.9 | 0.55 | **0.578** |
| **Time*Group[Location]** | 6 | 89.6 | 0.64 | **0.691** |
| **Age** | 1 | 51.3 | 0.89 | **0.349** |
| **Sex** | 1 | 52.5 | 0.19 | **0.659** |
| **NIHSS** | 1 | 55.6 | 1.09 | **0.299** |
| **MVC** | **1** | **51.8** | **2.91** | **0.094** |

CSP, cortical silent period; ICF, intracortical facilitation; MEP, motor evoked potential; MVC, maximal voluntary contraction; NIHSS, national institutes of health stroke scale; SICI, short-intracortical inhibition. Location represents the three lesion location groups—cortical, subcortical, and cerebellar—nested in the model as a categorical variable. * $p<0.05$

**Supplementary Table 7.** Adjusted multivariate linear regression examining associations between chronic CSE responses and changes in clinical motor outcomes and cardiorespiratory fitness for both groups.

| **Predictor** | **CE+standard care** | | | **Standard care** | | |
|---|---|---|---|---|---|---|
| | **Estimate (95% CI)** | **p value** | **$R^2$** | **Estimate (95% CI)** | **p value** | **$R^2$** |
| **UL-FMA T0-T2** | | | | | | |
| **Ipsilesional** | | | | | | |
| **Δ RMT** | -0.06 (-0.32, 0.19) | 0.596 | 0.27 | 0.03 (-0.30, 0.38) | 0.817 | 0.57 |
| **Δ Resting MEP** | 0.18 (-3.18, 3.56) | 0.910 | 0.28 | 3.94 (-4.03, 11.93) | 0.299 | 0.61 |
| **Δ Active MEP** | 0.93 (-1.54, 3.41) | 0.445 | 0.29 | -0.02 (-4.04, 4.00) | 0.990 | 0.57 |
| **Δ CSP** | -3.29 (57.09, 50.50) | 0.901 | 0.28 | -53.58 (-135.04, 27.87) | 0.175 | 0.64 |
| **Δ ICF** | 0.10 (-0.39, 0.60) | 0.662 | 0.28 | -0.37 (-1.31, 0.57) | 0.403 | 0.59 |
| **Δ SICI** | 0.94 (-0.54, 2.44) | 0.202 | 0.13 | -0.65 (-2.32, 1.02) | 0.408 | 0.59 |
| **Contralesional** | | | | | | |
| **Δ RMT** | -0.35 (-0.90, 0.19) | 0.194 | 0.32 | 0.10 (-0.13, 0.34) | 0.363 | 0.53 |
| **Δ Resting MEP** | 3.06 (-3.62, 9.76) | 0.357 | 0.30 | -3.58 (-11.50, 4.32) | 0.345 | 0.60 |
| **Δ Active MEP** | 1.79 (-0.05, 3.64) | 0.056 | 0.36 | 0.70 (-1.71, 3.11) | 0.541 | 0.59 |
| **Δ CSP** | -15.13 (-126.89, 96.63) | 0.784 | 0.28 | -8.36 (-76.33, 59.61) | 0.794 | 0.57 |
| **Δ ICF** | 0.99 (-0.33, 2.27) | 0.141 | 0.33 | 0.30 (-0.32, 0.94) | 0.313 | 0.61 |
| **Δ SICI** | 0.96 (-1.06, 2.99) | 0.336 | 0.17 | 0.59 (-1.05, 2.23) | 0.450 | 0.59 |
| **BBT T0-T2** | | | | | | |
| **Ipsilesional** | | | | | | |
| **Δ RMT** | 0.09 (-0.33, 0.52) | 0.659 | 0.03 | 0.30 (-0.34, 0.96) | 0.320 | 0.50 |
| **Δ Resting MEP** | 4.22 (-1.22, 9.68) | 0.123 | 0.10 | -1.72 (-16.79, 13.33) | 0.805 | 0.56 |
| **Δ Active MEP** | 0.53 (-3.68, 4.75) | 0.796 | 0.01 | -0.006 (-7.23, 7.22) | 0.998 | 0.55 |
| **Δ CSP** | -40.62 (-130.18, 48.93) | 0.360 | 0.05 | -4.16 (-163.93, 155.59) | 0.955 | 0.55 |
| **Δ ICF** | -0.47 (-1.29, 0.35) | 0.249 | 0.06 | 0.89 (-0.75, 2.54) | 0.256 | 0.61 |
| **Δ SICI** | -2.54 (-6.78, 1.70) | 0.226 | 0.10 | 1.77 (-1.10, 4.65) | 0.202 | 0.62 |
| **Contralesional** | | | | | | |
| **Δ RMT** | -0.19 (-1.06, 0.68) | 0.657 | 0.03 | 0.49 (0.05, 0.94) | 0.031* | 0.65 |

| | | | | | | |
|---|---|---|---|---|---|---|
| **Δ Resting MEP** | -3.89 (-14.37, 6.58) | 0.454 | 0.04 | 2.40 (-13.64, 18.46) | 0.751 | 0.51 |
| **Δ Active MEP** | -1.78 (-4.85, 1.27) | 0.243 | 0.07 | -1.89 (-6.56, 2.78) | 0.397 | 0.53 |
| **Δ CSP** | 35.11 (-140.77, 210.99) | 0.687 | 0.03 | 1.80 (-132.09, 135.70) | 0.977 | 0.51 |
| **Δ ICF** | -0.80 (-2.80, 1.19) | 0.417 | 0.06 | -0.90 (-2.08, 0.28) | 0.124 | 0.59 |
| **Δ SICI** | -2.60 (-6.53, 1.31) | 0.181 | 0.12 | -1.77 (-4.89, 1.35) | 0.243 | 0.56 |
| **CRF T0-T2** | | | | | | |
| **Ipsilesional** | | | | | | |
| **Δ RMT** | 0.007 (-0.14, 0.17) | 0.921 | 0.29 | 0.15 (-0.18, 0.48) | 0.341 | 0.28 |
| **Δ Resting MEP** | 2.47 (0.46, 4.49) | 0.017* | 0.39 | -4.59 (-12.68, 1.50) | 0.237 | 0.31 |
| **Δ Active MEP** | 0.77 (-0.86, 2.41) | 0.341 | 0.28 | 2.04 (-1.87, 5.95) | 0.275 | 0.30 |
| **Δ CSP** | -11.60 (-47.08, 23.87) | 0.508 | 0.27 | 29.83 (-59.44, 119.12) | 0.477 | 0.25 |
| **Δ ICF** | -0.34 (-0.64, 0.03) | 0.028* | 0.38 | -0.22 (-1.21, 0.77) | 0.633 | 0.23 |
| **Δ SICI** | 0.03 (-1.70, 1.78) | 0.963 | 0.30 | 0.30 (-1.46, 2.06) | 0.715 | 0.22 |
| **Contralesional** | | | | | | |
| **Δ RMT** | -0.52 (-0.81, -0.23) | 0.0008* | 0.46 | 0.08 (-0.14, 0.32) | 0.444 | 0.27 |
| **Δ Resting MEP** | -0.16 (-4.26, 3.92) | 0.933 | 0.23 | 2.62 (-5.40, 10.65) | 0.492 | 0.27 |
| **Δ Active MEP** | -1.19 (-1.37, 0.98) | 0.740 | 0.23 | -1.41 (-3.70, 0.87) | 0.204 | 0.33 |
| **Δ CSP** | -19.30 (-87.68, 49.07) | 0.569 | 0.24 | 9.61 (-58.13, 77.37) | 0.763 | 0.25 |
| **Δ ICF** | -0.02 (-0.78, 0.73) | 0.942 | 0.23 | 0.23 (-0.41, 0.87) | 0.412 | 0.28 |
| **Δ SICI** | -0.28 (-1.85, 1.28) | 0.712 | 0.27 | -0.35 (-2.01, 1.31) | 0.654 | 0.25 |

BBT, Box and Blocks Test; CRF, cardiorespiratory fitness; CSP, cortical silent period; ICF, intracortical facilitation; MEP, motor evoked potential; RMT, resting motor threshold; SICI short-intracortical inhibition. UL-FMA, upper-limb Fugl-Meyer assessment.

**Supplementary Table 8.** Adjusted multivariate linear regression examining associations between acute CSE responses over time and changes in clinical motor outcomes and cardiorespiratory fitness for both groups.

| Predictor | CE+standard care | | | Standard care | | |
|---|---|---|---|---|---|---|
| | Estimate (95% CI) | p value | $R^2$ | Estimate (95% CI) | p value | $R^2$ |
| **UL-FMA T0-T2** | | | | | | |
| **Ipsilesional** | | | | | | |
| **Δ Resting MEP** | -1.02 (-4.99, 2.94) | 0.602 | 0.29 | -2.44 (-9.10, 4.22) | 0.436 | 0.59 |
| **Δ Active MEP** | 0.49 (-3.17, 4.17) | 0.784 | 0.28 | -1.13 (-5.32, 3.06) | 0.564 | 0.58 |
| **Δ CSP** | 12.18 (-36.98, 61.36) | 0.616 | 0.28 | -6.29 (-92.18, 79.60) | 0.874 | 0.57 |
| **Δ ICF** | 0.30 (-0.31, 0.93) | 0.324 | 0.31 | 0.15 (-0.48, 0.78) | 0.609 | 0.58 |
| **Δ SICI** | -0.32 (-1.45, 0.80) | 0.559 | 0.08 | 0.10 (-1.28, 1.50) | 0.866 | 0.57 |
| **Contralesional** | | | | | | |
| **Δ Resting MEP** | -1.11 (-6.33, 4.10) | 0.666 | 0.29 | 0.91 (-4.31, 6.14) | 0.710 | 0.58 |
| **Δ Active MEP** | 0.41 (-2.03, 2.85) | 0.734 | 0.31 | 0.98 (-2.29, 4.26) | 0.528 | 0.59 |
| **Δ CSP** | -22.51 (-129.04, 84.02) | 0.669 | 0.31 | 14.83 (-39.25, 68.93) | 0.563 | 0.58 |
| **Δ ICF** | -0.09 (-0.78, 0.59) | 0.788 | 0.28 | 0.07 (-0.42, 0.57) | 0.755 | 0.58 |
| **Δ SICI** | -0.69 (-2.03, 0.64) | 0.293 | 0.17 | -0.43 (-1.95, 1.08) | 0.293 | 0.17 |
| **BBT T0-T2** | | | | | | |
| **Ipsilesional** | | | | | | |
| **Δ Resting MEP** | -4.91 (-11.46, 1.62) | 0.135 | 0.09 | -7.52 (-18.80, 3.74) | 0.169 | 0.63 |
| **Δ Active MEP** | -3.53 (-9.58, 2.52) | 0.242 | 0.06 | -2.31 (-9.82, 5.19) | 0.511 | 0.57 |
| **Δ CSP** | -11.71 (-44.06, 20.62) | 0.464 | 0.27 | -19.75 (-173.79, 134.27) | 0.782 | 0.56 |
| **Δ ICF** | 0.50 (-0.55, 1.57) | 0.337 | 0.05 | 0.008 (-1.14, 1.15) | 0.987 | 0.55 |
| **Δ SICI** | 2.52 (-0.48, 5.54) | 0.095 | 0.16 | -0.30 (-2.80, 2.19) | 0.793 | 0.56 |
| **Contralesional** | | | | | | |
| **Δ Resting MEP** | -7.07 (-14.79, 0.64) | 0.071 | 0.12 | -7.18 (-16.57, 2.20) | 0.122 | 0.59 |
| **Δ Active MEP** | 2.76 (-1.12, 6.64) | 0.157 | 0.08 | 2.29 (-4.10, 8.69) | 0.452 | 0.53 |
| **Δ CSP** | 113.88 (-50.60, 278.38) | 0.167 | 0.08 | 56.87 (-45.28, 159.03) | 0.250 | 0.56 |
| **Δ ICF** | 1.11 (0.11, 2.10) | 0.059* | 0.16 | 0.69 (-0.19, 1.58) | 0.113 | 0.60 |

| | | | | | | |
|---|---|---|---|---|---|---|
| **Δ SICI** | 1.76 (-1.03, 4.57) | 0.204 | 0.08 | 2.45 (-0.10, 5.10) | 0.067 | 0.62 |
| **CRF T0-T2** | | | | | | |
| **Ipsilesional** | | | | | | |
| **Δ Resting MEP** | -2.52 (-5.00, -0.04) | 0.046* | 0.36 | 5.52 (-0.50, 11.55) | 0.068 | 0.42 |
| **Δ Active MEP** | -1.66 (-4.03, 0.69) | 0.159 | 0.31 | -3.99 (-7.48, -0.50) | 0.028* | 0.50 |
| **Δ CSP** | -11.71 (-44.06, 20.62) | 0.464 | 0.27 | -77.57 (-149.53, -5.60) | 0.037* | 0.48 |
| **Δ ICF** | 0.28 (-0.11, 0.69) | 0.156 | 0.30 | 0.06 (-0.59, 0.72) | 0.827 | 0.21 |
| **Δ SICI** | 0.24 (-1.03, 1.52) | 0.696 | 0.31 | -0.30 (-1.72, 1.11) | 0.641 | 0.23 |
| **Contralesional** | | | | | | |
| **Δ Resting MEP** | -0.99 (-4.25, 2.26) | 0.538 | 0.24 | 2.92 (-2.01, 7.87) | 0.223 | 0.33 |
| **Δ Active MEP** | -0.31 (-1.86, 1.23) | 0.682 | 0.24 | -0.08 (-3.41, 3.23) | 0.956 | 0.24 |
| **Δ CSP** | 14.76 (-54.31, 83.85) | 0.665 | 0.24 | -21.87 (-74.97, 31.22) | 0.389 | 0.29 |
| **Δ ICF** | 0.46 (0.07, 0.85) | 0.021* | 0.35 | -0.27 (-0.74, 0.20) | 0.238 | 0.32 |
| **Δ SICI** | 0.02 (-1.16, 1.20) | 0.968 | 0.28 | 0.71 (-0.76, 2.19) | 0.314 | 0.30 |

BBT, Box and Blocks Test; CRF, cardiorespiratory fitness; CSP, cortical silent period; ICF, intracortical facilitation; MEP, motor evoked potential; SICI short-intracortical inhibition. UL-FMA, upper-limb Fugl-Meyer assessment.
